# Alterations of the upper respiratory microbiome among children living with HIV infection in Botswana

**DOI:** 10.1101/2022.12.19.22283664

**Authors:** Sweta M. Patel, John Farirai, Mohamed Z. Patel, Sefelani Boiditswe, Leabaneng Tawe, Shimane Lekalake, Mogomotsi Matshaba, Andrew P. Steenhoff, Tonya Arscott-Mills, Kristen A. Feemster, Samir S. Shah, Nathan Thielman, Coleen K. Cunningham, Lawrence A. David, David M. Murdoch, Matthew S. Kelly

## Abstract

Children living with HIV (CLWH) are at high risk of colonization and infection by bacterial respiratory pathogens, though this risk can be reduced by other microbes in the upper respiratory microbiome. The impact of HIV infection on development of the upper respiratory microbiome during childhood is poorly understood, and we sought to address this knowledge gap by identifying associations between HIV infection and the nasopharyngeal microbiomes of children in Botswana. We enrolled Batswana CLWH (<5 years) and age- and sex-matched HIV-exposed, uninfected (HEU) and HIV-unexposed, uninfected (HUU) children in a cross-sectional study. We used shotgun metagenomic sequencing to compare the nasopharyngeal microbiomes of children by HIV status. Among the 143 children in this study, HIV infection and HIV-associated immunosuppression were associated with alterations in nasopharyngeal microbiome composition, including lower abundances of *Corynebacterium* species associated with respiratory health. These findings suggest that the upper respiratory microbiome may contribute to the high risk of bacterial respiratory infections among CLWH.

## INTRODUCTION

Despite widespread use of the 13-valent pneumococcal conjugate (PCV-13) and *H. influenzae* type B (Hib) vaccines, bacterial respiratory pathogens including *Streptococcus pneumoniae* (pneumococcus) and *Staphylococcus aureus* account for considerable child morbidity and mortality worldwide.^1,2^ The nasopharynx is an important pathogen reservoir, and nasopharyngeal colonization precedes the development of invasive infection.^3^ Colonization is influenced both by host-microbe interactions and interactions with other nasopharyngeal microbes. For example, *Corynebacterium accolens* hydrolyzes human triacylglycerols to produce free fatty acids that inhibit pneumococcal growth.^4^ Given the role that microbes in the nasopharynx play in resisting colonization and infections caused by bacterial respiratory pathogens, targeted manipulation of the nasopharyngeal microbiome is a promising strategy for the prevention or treatment of these infections.

The nasopharyngeal microbiome may represent a particularly important therapeutic target in children living with HIV (CLWH). CLWH have higher incidences of nasopharyngeal colonization and invasive infections caused by pneumococcus and *S. aureus*, likely resulting from multiple mechanisms.^5,6^ Maternal HIV infection impairs placental transfer of antibodies to respiratory pathogens including pneumococcus.^7^ Socioeconomic factors, replacement feeding, and undernutrition may further contribute but fail to fully explain the rates of infections observed in this population. Variations in the upper respiratory microbiome may also affect the risk of bacterial pathogen colonization and infection among CLWH. Prior studies demonstrated that the gut and oral microbiomes are altered in CLWH.^8,9^ However, few data exist on associations between HIV infection and the nasopharyngeal microbiome.^10^

In this study, we sought to identify associations between HIV infection and the nasopharyngeal microbiomes of children in Botswana. We used shotgun metagenomic sequencing to compare the nasopharyngeal microbiomes of CLWH, HIV-exposed, uninfected (HEU) children, and HIV-unexposed, uninfected (HUU) children. We identified HIV-related and environmental factors associated with nasopharyngeal microbiome composition during childhood.

## METHODS

### Setting

This study was conducted from February 2020 to February 2021 at five public clinics in Gaborone, Botswana. The country’s HIV prevalence among pregnant women is 24%.^11^ The government of Botswana provides antiretroviral therapy (ART) at no cost, and 98% of pregnant women receive ART for prevention of vertical HIV transmission.^12^ All children in Botswana are eligible to receive free childhood vaccinations at public clinics in Botswana and CLWH receive free ART. Botswana incorporated a pentavalent vaccine containing Hib (Serum Institute of India) into its immunization program in November 2010 and PCV-13 (Prevnar-13; Pfizer) in July 2012; both vaccines are administered in 3+0 schedules with doses at 2, 3, and 4 months of age. The estimated national coverage rates among infants in 2020 for the complete Hib and PCV-13 vaccine series were 95% and 90%, respectively.^13^

### Study Population

Children under five years of age with documentation of child and maternal HIV testing and without chronic pulmonary disease or clinical signs of pneumonia were eligible for inclusion. We excluded children with craniofacial abnormalities because of possible increased risk of injury from nasopharyngeal specimen collection. For each child living with HIV, we enrolled an HEU and an HUU child, each sex- and age-matched (±3 months). Additionally, we enrolled any HEU siblings under five years of age with the same biological mother who lived in the same household as enrolled CLWH. Written informed consent was obtained for all study participants from the accompanying parent or guardian. This study was approved by the Health Research and Development Committee (Ministry of Health, Botswana) and institutional review boards at the University of Botswana, the Botswana-Baylor Children’s Clinical Centre of Excellence, the University of Pennsylvania, and the Duke University Health

### Data collection

We collected data through physical examination, a questionnaire with the child’s guardian, and review of medical records. Children with a positive HIV PCR were classified as CLWH, while children born to women with HIV at or before delivery were classified as HEU if they had documentation of a negative HIV PCR after 6 weeks of age if exclusively formula fed or at least 6 weeks after breastfeeding cessation. Children born to mothers with negative testing for HIV during or after pregnancy were classified as HUU. Using the most recent results available at the time of enrollment, we defined viral suppression as an HIV RNA <400 copies/mL, a normal CD4+ cell percentage (CD4+%) as ≥25%, and a low CD4+% as <25%.^14^

### Laboratory testing (Supplemental File p2)

Nasopharyngeal specimens were collected using flocked swabs and MSwab medium (Copan Italia, Brescia, Italy). Specimens were stored at -80°C at the National Health Laboratory in Gaborone, Botswana before shipment to Duke University. The Duke University Microbiome Core Facility extracted DNA from nasopharyngeal specimens using DNeasy PowerSoil Pro kits (Qiagen, Hilden, Germany). Sequencing libraries were generated using DNA Prep kits (Illumina, San Diego, CA, USA). Libraries were pooled at equimolar concentrations and sequenced as 150 base pair paired-end reads on a NovaSeq6000 instrument (Illumina) in the Duke University Sequencing and Genomic Technologies core facility. Positive and negative extraction and library preparation controls were included on the sequencing run.

### Bioinformatics processing

Initial filtering and quality assessment of the sequencing data were performed using the default settings in the KneadData toolkit version 0.10.0, which filtered reads based on nucleotide quality, adapter content, contamination from human reads, and low-information tandem repeats using Trimmomatic v0.39 and Bowtie 2 v2.4.4.^15,16^ Kaiju version 1.8.2 was used to assign taxonomy to sequencing reads using the proGenomes database version 2·1 as the reference, 10 sequencing reads across the dataset were filtered, as were non-bacterial reads. We used the *decontam* R package version 1·14 to remove presumed reagent contaminants (n=53) based on negative correlation with DNA concentration using the frequency method (threshold=0·1; Table S1, pp 3-4). Based on rarefaction curves generated to determine the pruning threshold, samples with fewer than 2,500 high-quality paired-end reads were excluded from further analyses (Figure S1, p14).

### Statistical analysis (Supplemental File pp 1-2)

We used multivariable linear regression to evaluate associations between HIV status and the Shannon index and log-transformed Chao1 richness of the microbiome. We used permutational analysis of variance (PERMANOVA) to compare community-level microbiome composition by HIV status. We used paired PERMANOVA testing to compare community-level microbiome composition within sibling pairs. In addition to identifying differences in microbiome composition by HIV status, we used unsupervised k-medoids clustering and the Calinksi-Harabasz index to generate nasopharyngeal microbiome profiles and evaluated associations between HIV status and microbiome profiles using multinomial logistic regression. After filtering taxa with fewer than 50 counts across the dataset, we fit multivariable general linear models using MaAsLin 2·0 to identify differentially abundant bacterial species by HIV status.^19^ For analyses limited to CLWH, we filtered taxa as described above and used the linear discriminant effect size (LEfSe) method to identify differentially abundant species by CD4+%, viral suppression status, use of trimethoprim-sulfamethoxazole (TMP-SMX) prophylaxis, and antibiotic treatment in the preceding three months.^20^ We used Wilcoxon signed-rank tests to compare the relative abundances of the ten most common bacterial species within sibling pairs. Lastly, we used Sparse Correlations for Compositional Data (SparCC) to evaluate associations between the relative abundances of bacterial species. Unless otherwise noted, we adjusted all analyses for potential confounding variables identified based on a review of the literature: season, age, household use of wood for cooking or heating, receipt of antibiotics within the preceding three months, and upper respiratory infection (URI) symptoms (nasal congestion, nasal discharge, or cough) in the threshold for statistical significance was p<0.05 for analyses that did not adjust for multiple testing. The Benjamini-Hochberg procedure was used to adjust for multiple testing in multivariable general linear models and a threshold of padj (also known as q) <0.2 was considered statistically significant. Similarly, the Benjamini-Hochberg procedure was used to adjust for multiple testing for analyses conducted using SparCC and a threshold of padj<0.1 was considered statistically significant.

## RESULTS

### Characteristics of the study population

Of 187 children screened for eligibility, 153 were enrolled (Figure 1). Sufficient microbial reads for analysis were obtained from the nasopharyngeal samples of 143 (93%) children, including 44 (31%) CLWH, 49 (34%) HEU, and 50 (35%) HUU (Table 1). The median (interquartile range, IQR) number of raw reads per sample was 38,813,926 (33,294,776; 43,851,661), >99% of which were human in origin. The median (IQR) number of microbial reads per sample was 77,097 (18,847-155,456); sample sequencing depth did not differ by HIV exposure status (Kruskal-Wallis test; p=0.55). Forty-three of 44 CLWH were enrolled from HIV subspecialty clinics and all HEU and HUU children were enrolled from three general child health clinics. Median (IQR) age was 32 (18, 49) months, all enrolled children were Black African, and 52% of children were female. Maternal age (p=0·0002), breastfeeding history (p<0·0001), and height-for-age (p=0·0005) and weight-for-age (p<0·0001) z-scores differed by child HIV status. Of CLWH, 42 (95%) were prescribed ART with the most frequent regimen being abacavir-lamivudine-lopinavir/ritonavir (n=38). Median (IQR) CD4+% was 30% (24-36%), and 27 of 41 (66%) children with HIV RNA measurements had undetectable viral loads (<400 copies/mL) collected a median (IQR) of 12 (4, 18) weeks prior to enrollment. Eleven of 42 children (26%) had a low CD4+%.

**Figure 1.**
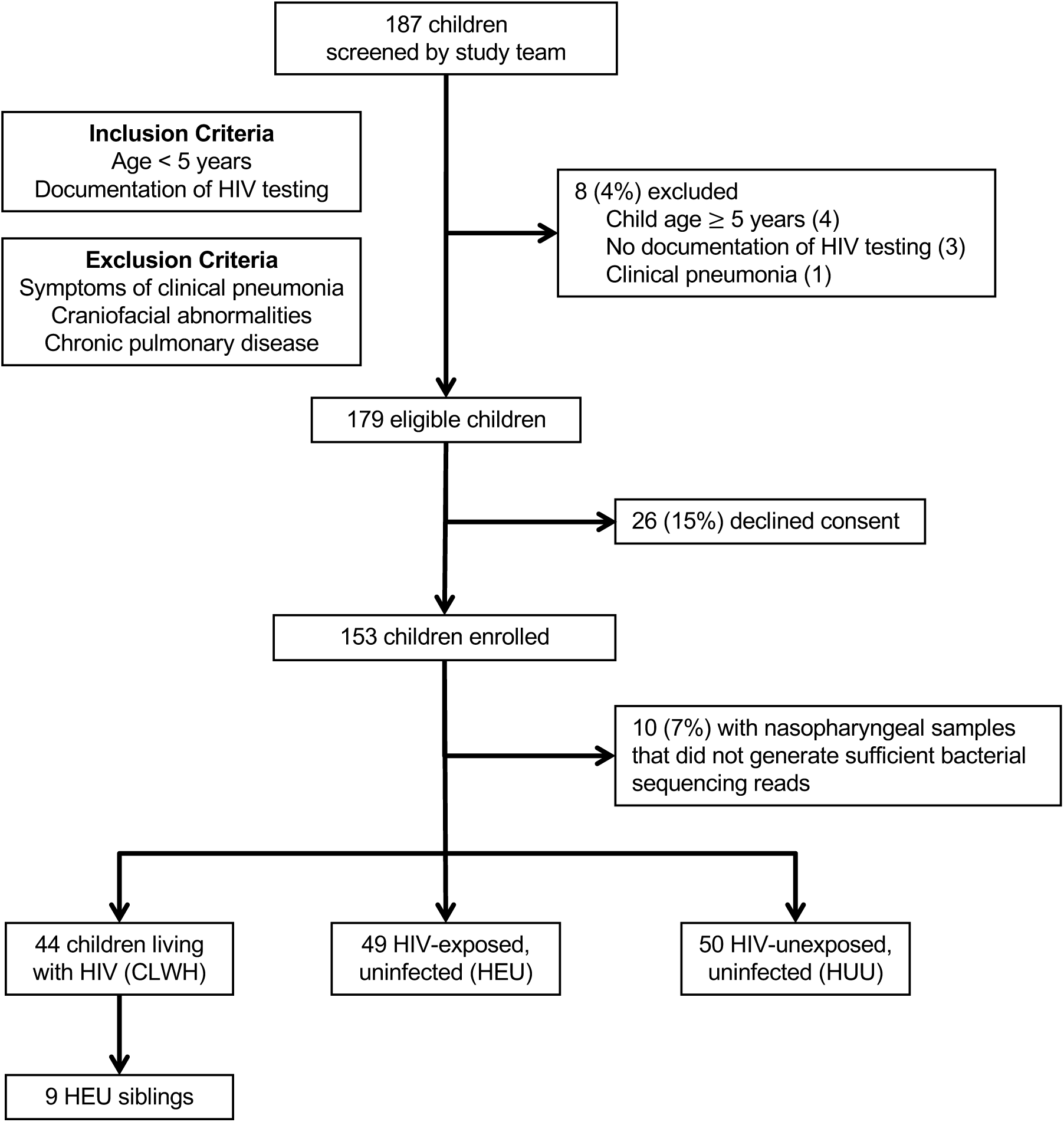
Screening and enrollment of children in Gaborone, Botswana, February 2020 to February 2021.

**Table 1.** Baseline characteristics of the study population by HIV status.

|  |  | HIV Status |  |  |  |
| --- | --- | --- | --- | --- | --- |
|  | Overall<br>(n=143) | CLWH<br>(n=44) | HEU<br>Children<br>(n=49) | HUU<br>Children<br>(n=50) | p* |
|  | N (%) | N (%) | N (%) | N (%) |  |
| **Demographics** |  |  |  |  |  |
| Child age in months, median (IQR) | 32 (18, 49) | 30 (19, 45) | 32 (17, 49) | 35 (18, 49) | 0·83 |
| Female sex | 74 (52) | 23 (52) | 25 (51) | 26 (52) | 0·99 |
| Maternal age, y, median (IQR) | 29 (25, 35) | 30 (25, 35) | 34 (27, 39) | 26 (24, 30) | 0·0002 |
| Rainy season (November to March) | 90 (63) | 29 (66) | 30 (61) | 31 (62) | 0·88 |
| **Socioeconomic factors** |  |  |  |  |  |
| Maternal education level (N=142) |  |  |  |  | 0·38 |
| None or primary | 15 (11) | 6 (14) | 7 (14) | 2 (4) |  |
| Secondary | 110 (77) | 31 (72) | 37 (76) | 42 (84) |  |
| Tertiary | 17 (12) | 6 (14) | 5 (10) | 6 (12) |  |
| Electricity in home | 106 (74) | 35 (80) | 32 (65) | 39 (78) | 0·22 |
| Use of wood for cooking or heating | 87 (61) | 26 (59) | 32 (65) | 29 (58) | 0·73 |
| Number of household members, median (IQR) | 4 (3,6) | 5 (3,7) | 4 (3,6) | 4 (3,5) | 0·51 |
| **Clinical factors** |  |  |  |  |  |
| Antibiotic exposure in prior 3 months | 24 (17) | 7 (16) | 10 (20) | 7 (14) | 0·68 |
| URI symptoms in prior 1 month | 59 (41) | 20 (45) | 19 (39) | 20 (40) | 0·79 |
| Received 3 doses of PCV-13 (N=141) | 127 (90) | 37 (88) | 44 (90) | 46 (92) | 0·31 |
| Received 3 doses of Hib vaccine (N=141) | 130 (92) | 36 (86) | 46 (94) | 48 (96) | 0·50 |
| Ever breastfed | 84 (59) | 21 (48) | 16 (33) | 47 (94) | <0·0001 |
| Weight-for-age z-scores, median (IQR) | -0·31 (-1·11, 0·20) | -1·00 (-1·53, -0·33) | -0·10 (-0·64, 0·58) | -0·09 (-0·91, 0·14) | <0·0001 |
| Height-for-age z-scores, median (IQR) | -0·32 (-1·29, 0·43) | -1·00 (-1·93, -0·14) | -0·10 (-0·94, 0·44) | -0·07 (-0·83, 0·64) | 0·0005 |

CLWH, children living with HIV; HEU, HIV-exposed, uninfected; HUU, HIV-unexposed, uninfected; IQR, interquartile range; URI, upper respiratory infection; PCV-13, 13-valent pneumococcal conjugate vaccine; Hib, *H. influenzae* type B
\*p values were estimated using Chi-square or Fisher's exact tests for categorical variables and Kruskal-Wallis tests for continuous variables

Fifteen (36%) CLWH were receiving trimethoprim-sulfamethoxazole (TMP-SMX) prophylaxis. Seven (16%) had received other antibiotic treatment in the preceding 3 months.

### HIV infection is associated with community-level differences in the nasopharyngeal microbiome

A total of 6,709 bacterial species from 116 phyla and 2,146 genera were detected, with Actinobacteria, Firmicutes, and Proteobacteria being the most abundant phyla (Table S2, p5). Median (IQR) Shannon and Chao1 indices were 1·85 (1·51, 2·26) and 137 (68, 261), respectively. Using multivariable linear regression, there were no associations between HIV status and measures of alpha diversity (Table S3, p6; Figure S2, p15). The community-level composition of the nasopharyngeal microbiome differed by HIV status (Figure 2A, PERMANOVA; p=0·043, R^2^=0·019), as well as by child age, season of enrollment, and recent URI symptoms (Table S4, p7). To further investigate the observed association between HIV and microbiome composition, we used unsupervised clustering to classify nasopharyngeal samples into five distinct microbiome profiles (Figure 3) and evaluated associations between these microbiome profiles and HIV status. Participant age (p=0·042), season of enrollment (p=0·0078), household size (p=0·0022), and recent URI symptoms (p=0·033) differed by microbiome profile (Table S5, p8). In adjusted analyses, CLWH were more likely to have the biodiverse (BIO) profile, which also contained a higher relative abundance of pneumococcus than the other microbiome profiles (multinomial logistic regression; relative risk ratio (95% confidence interval (CI)): 5·54 (1·37, 22·3); Table S6, p9).

**Figure 2.**
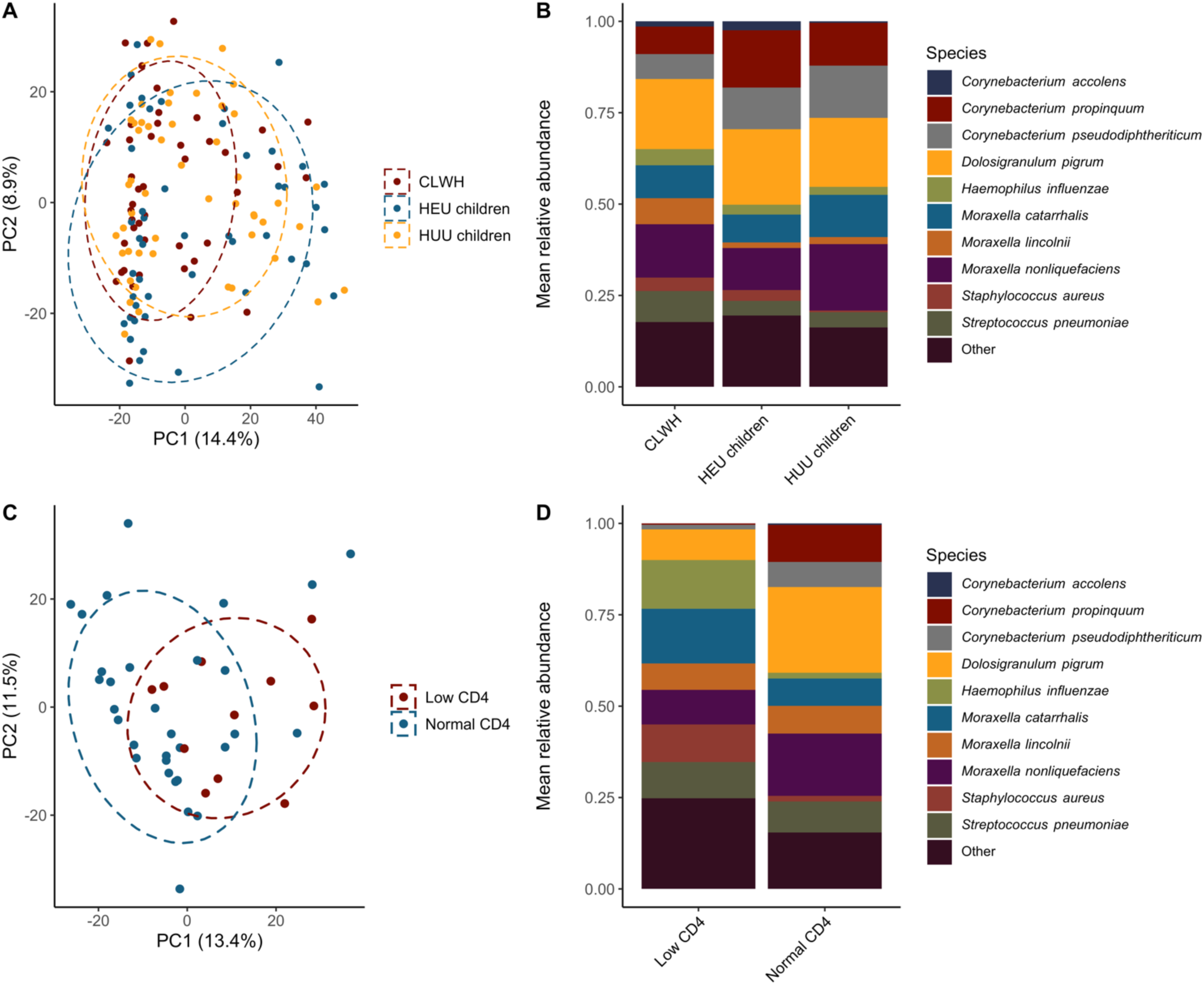
Composition of the nasopharyngeal microbiome by HIV status and HIV-associated immunosuppression among children in Botswana. A. Principal components plot based on Euclidean distances showing distinct nasopharyngeal microbiome composition by HIV status among children in Botswana (n=143, PERMANOVA: p = 0·043, R^2^ = 0·019). B. Relative abundances of the ten most abundant bacterial species by HIV status in nasopharyngeal samples from children in Botswana (n=143). C. Principal components plot based on Euclidean distances showing distinct nasopharyngeal microbiome composition by CD4+ cell percentage among children living with HIV (CLWH) in Botswana (n=44, PERMANOVA: p=0·009, R^2^ = 0·042). D. Relative abundances of the ten most abundant bacterial species by CD4+ cell percentage in nasopharyngeal samples from CLWH in Botswana (n=44).

**Figure 3.**
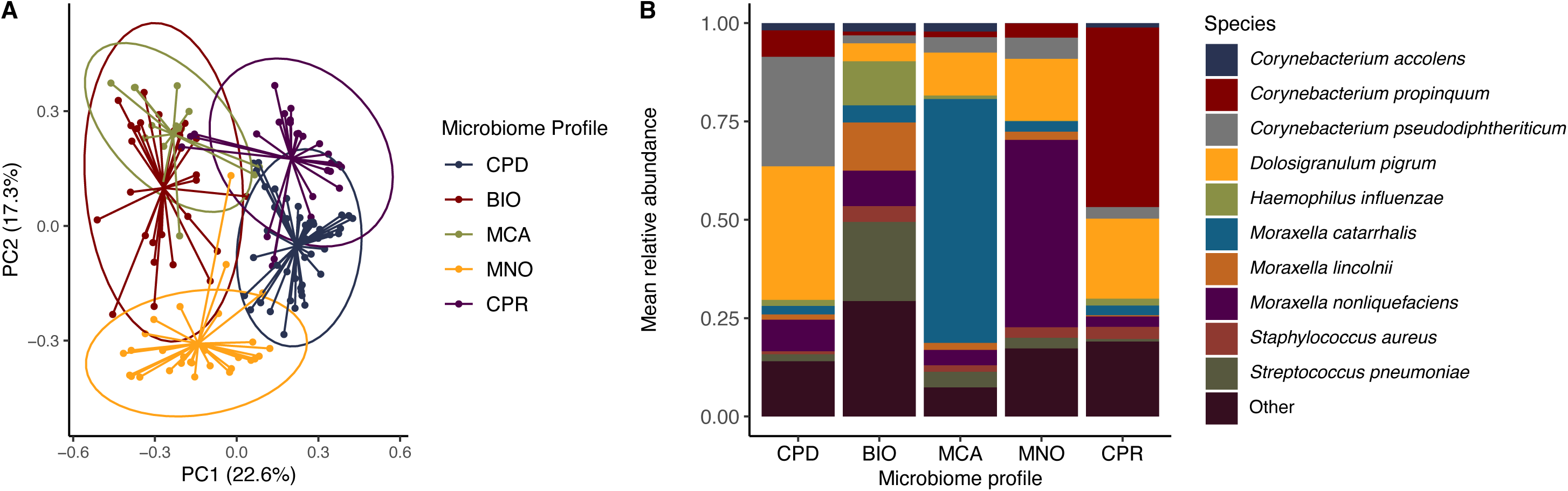
Nasopharyngeal microbiome profiles among children in Botswana generated by unsupervised clustering. K-medoids clustering and the Calinksi-Harabasz index were used to generate five nasopharyngeal microbiome profiles. A. Principal components plot based on Bray-Curtis distances showing clustering of nasopharyngeal samples by microbiome profile. Each dot represents one nasopharyngeal sample. Centroids are displayed as the confluence of lines arising from samples in each microbiome profile. Ellipses define the regions containing 95% of all samples that can be drawn from the underlying multivariate t distribution. B. Relative abundances of the ten most abundant bacterial species by microbiome profile. CPD, *C. pseudodiphtheriticum* and *D. pigrum*-dominated. BIO, biodiverse profile not dominated by a single species. MCA, *M. catarrhalis*-dominated. MNO, *M. nonliquefaciens*-dominated. CPR, *C. propinquum*-dominated.

### Certain species are differentially abundant in HIV infection

To further characterize compositional differences in the nasopharyngeal microbiome, we used MaAsLin2 to fit multivariable general linear models evaluating associations between sociodemographic and environmental factors and the abundances of bacterial species in the nasopharyngeal microbiome (Figure 4, Tables S7-S8, pp 10-11). In a model comparing CLWH to HUU children, the relative 2·22 (-3·88, -0·56); q=0·15). In a model comparing CLWH to HEU children, the relative abundances of *Corynebacterium accolens* (effect estimate (95% CI): -2·06 (-3·22, -0·92); q=0·091) and *Corynebacterium aurimucosum* (effect estimate (95% CI): -2·07 (-3·53, -0·61); q=0·016) were lower in CLWH (Figure 2B, Figure 4B). There were no differences in species abundances between HEU and HUU children. The relative abundance of *Micrococcus luteus* was higher and the relative abundances of multiple *Moraxella* species lower among children enrolled during the rainy season (Figure 4A). In contrast, recent URI symptoms were associated with a higher relative abundance of *Moraxella cuniculi* (Figure 4C). Finally, increasing age was associated with lower relative abundances of multiple non-pneumococcal *Streptococcus* species and higher relative abundances of *M. luteus* (Figure 4D).

**Figure 4.**
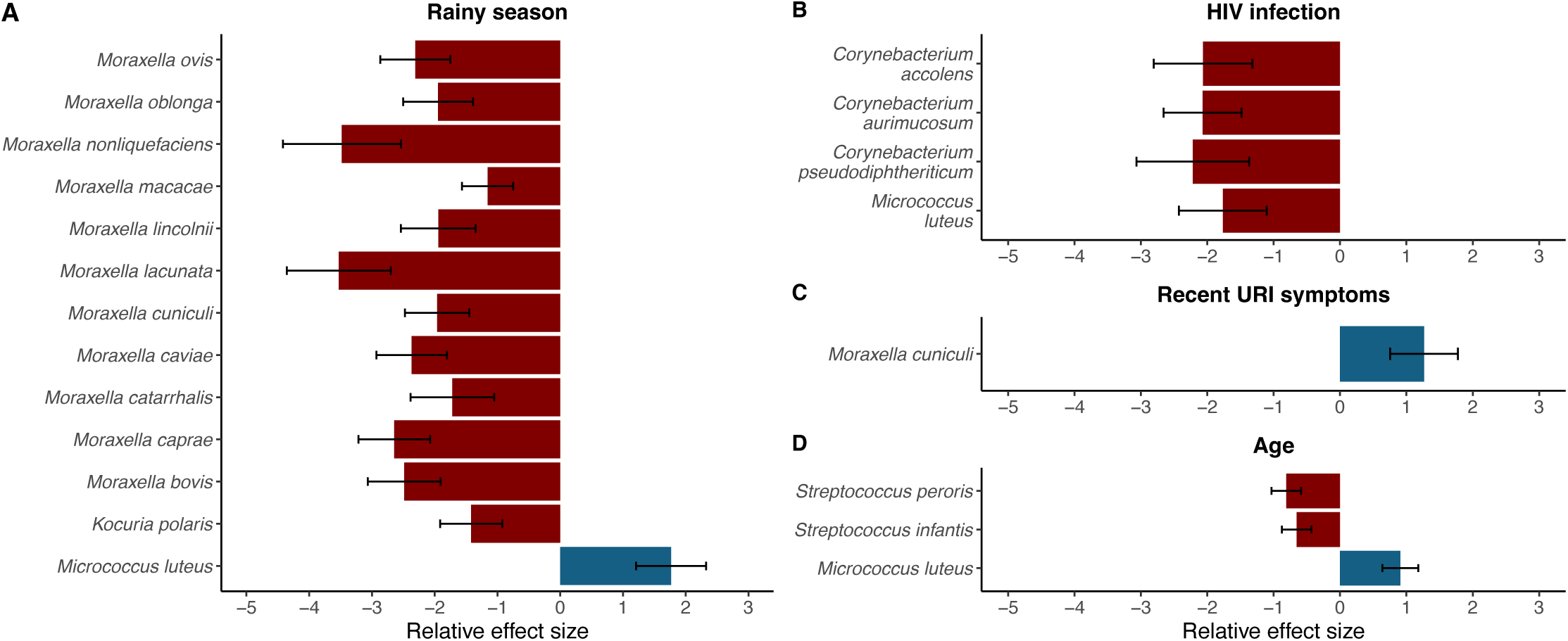
Associations between sociodemographic and clinical factors and environmental exposures and the composition of the microbiome among children in Botswana. MaAsLin2 was used to fit log-transformed generalized linear mixed models evaluating associations between subject factors and the relative abundances of bacterial species within the nasopharyngeal microbiome. The coefficients from these models, which correspond to the relative effect sizes of associations, are shown for significant associations (q < 0·20) identified for (A) rainy season, (B) HIV infection, (C) recent URI symptoms, and (D) age. (B) displays species that were differentially abundant in CLWH compared to HUU children (*C. pseudodiphtheriticum*) and species that were differentially abundant in CLWH compared to HEU children (*C. accolens, C. aurimucosum, M. luteus*). Bacterial species that were higher in relative abundance with the exposure are shown as blue bars. Bacterial species that were lower in relative abundance with the exposure are shown as red bars. Standard errors are represented by the error bars. URI, upper respiratory symptoms.

Given the significant difference in breastfeeding practices by HIV status and the previously established association between breastfeeding and microbiome composition, we performed a sensitivity analysis to evaluate the extent to which breastfeeding may contribute to the observed differences in microbiome composition by HIV status. In an analysis restricted to children with a history of breastfeeding (n=84), community-level differences in nasopharyngeal microbiome composition were observed by HIV status (Figure S3, p16, PERMANOVA; p=0·01, R^2^=0·036) and the relative abundances of *Corynebacterium accolens* (general linear model; effect size (95% CI): -4·06 (-1·96, -6·16); q=0·030) and *Corynebacterium aurimucosum* (effect size (95% CI): -3·10 (-1·26, -4·94); q=0·079) were lower in CLWH compared to HEU children (Table S9, p12).

We additionally compared the nasopharyngeal microbiomes of CLWH to their HEU siblings to more precisely identify associations between HIV infection and microbiome composition, given that environmental exposures among sibling pairs in the same household are likely to be similar. Among the nine sibling pairs with sufficient bacterial sequencing reads for analysis, there were no differences in demographic characteristics between CLWH and their HEU siblings (Table S10, p13); however, Additionally, community-level nasopharyngeal microbiome composition did not differ by HIV status (paired PERMANOVA; p=0·20, R^2^=0·075). However, CLWH had lower relative abundances of *Corynebacterium propinquum* (p=0·0078), *C. pseudodiphtheriticum* (p=0·0078), and *Dolosigranulum pigrum* (p=0·027), and higher relative abundances of *Moraxella lincolnii* (p=0·0039) and pneumococcus (p=0·021) than their HEU siblings (Wilcoxon signed-rank test; Figure S4, p17).

### HIV-associated immunosuppression is associated with altered nasopharyngeal microbiome composition among CLWH

We next sought to identify factors associated with the diversity and composition of the nasopharyngeal microbiome among CLWH. Using multivariable linear regression, there were no associations between viral suppression status, CD4+%, TMP-SMX exposure, or antibiotic treatment and the alpha diversity of the nasopharyngeal microbiome. Community-level microbiome composition differed between children with low and normal CD4+% (Figures 2C and 2D; PERMANOVA; p=0·013, R^2^=0·042), but did not differ by viral suppression status (p=0·79, R^2^=0·02), TMP-SMX exposure (p=0·13, R^2^=0·031), or recent antibiotic treatment (p=0·19, R^2^=0·029). In differential abundance analyses, *C. propinquum (*linear discriminant analysis (LDA) score 5·02)*, C. pseudodiphtheriticum* (LDA score 4·80)*, D. pigrum* (LDA score 5·25), and *Alloiococcus otitis* (LDA score 3·42) were more abundant in children with normal CD4+% compared to children with low CD4+%, and *Neisseria lactamica* (LDA score 4·17) was more abundant in children with low CD4+% (Figure S5A, p18; LEfSe). TMP-SMX exposure was associated with a higher relative abundance of *S. aureus* (LDA score 4·99, Figure S5B, p18). There were no differentially abundant species by viral suppression status or recent antibiotic exposure.

To further evaluate the association between HIV-associated immunosuppression and nasopharyngeal microbiome composition, we compared CLWH and low CD4+% to HEU and HUU children. Community-level microbiome composition differed in CLWH and low CD4+% compared to HEU and HUU children 4·46) were more abundant in HEU children compared to HUU children and immunosuppressed CLWH, while *C. pseudodiphtheriticum* (LDA score 5·00) and *C. aurimucosum* (LDA score 4·11) were more abundant in HUU children compared to HEU children and immunosuppressed CLWH (Figure S5C, p18; LEfSe). In contrast, community-level microbiome composition (PERMANOVA; p=0·15, R^2^=0·018) and individual species relative abundances did not differ between CLWH with normal CD4+% and HEU and HUU children (Figure S6, p19).

We also examined associations between microbiome profiles and viral suppression and immune status among CLWH. The proportion of CLWH with viral suppression was highest in the two profiles dominated by *C. pseudodiphtheriticum/D. pigrum* (CPD) or *M. nonliquefaciens* (MNO) compared to the other three profiles (85% vs 48%, p=0.020). The proportion of CLWH with low CD4+% was highest in the biodiverse (BIO) or *M. catarrhalis*-dominated (MCA) profiles when compared to the other three profiles combined (50% vs 8%, p=0.0040).

### Identification of species negatively associated with *S. pneumoniae* and *S. aureus* abundance

Finally, we sought to determine if differences in pneumococcal and *S. aureus* colonization existed by HIV status and how the abundances of other species were associated with the prevalence and abundance of these pathogens. Pneumococcus was identified in 126 (88%) children and *S. aureus* was identified in 45 (31%) children. There were no differences in pneumococcal colonization prevalence (Chi-squared test; p=0·094) or relative abundance (Kruskal-Wallis test; p=0·55) by HIV status. Conversely, S. aureus colonization was more common among HEU children than CLWH or HUU children (p=0·027), though the relative abundance did not differ by HIV status (p=0·049). Given that other microbes in the nasopharynx contribute to colonization resistance to pneumococcus and *S. aureus*, we evaluated correlations between the relative abundances of these potential pathogens and the abundances of the most abundant bacterial species in the nasopharyngeal microbiome using relative abundances of *C. propinquum, C. pseudodiphtheriticum, and D. pigrum* (Figure 5, Figure S7, p20), the latter of which are species whose abundances were also noted to be lower in the setting of HIV or HIV-associated immunosuppression.

**Figure 5.**
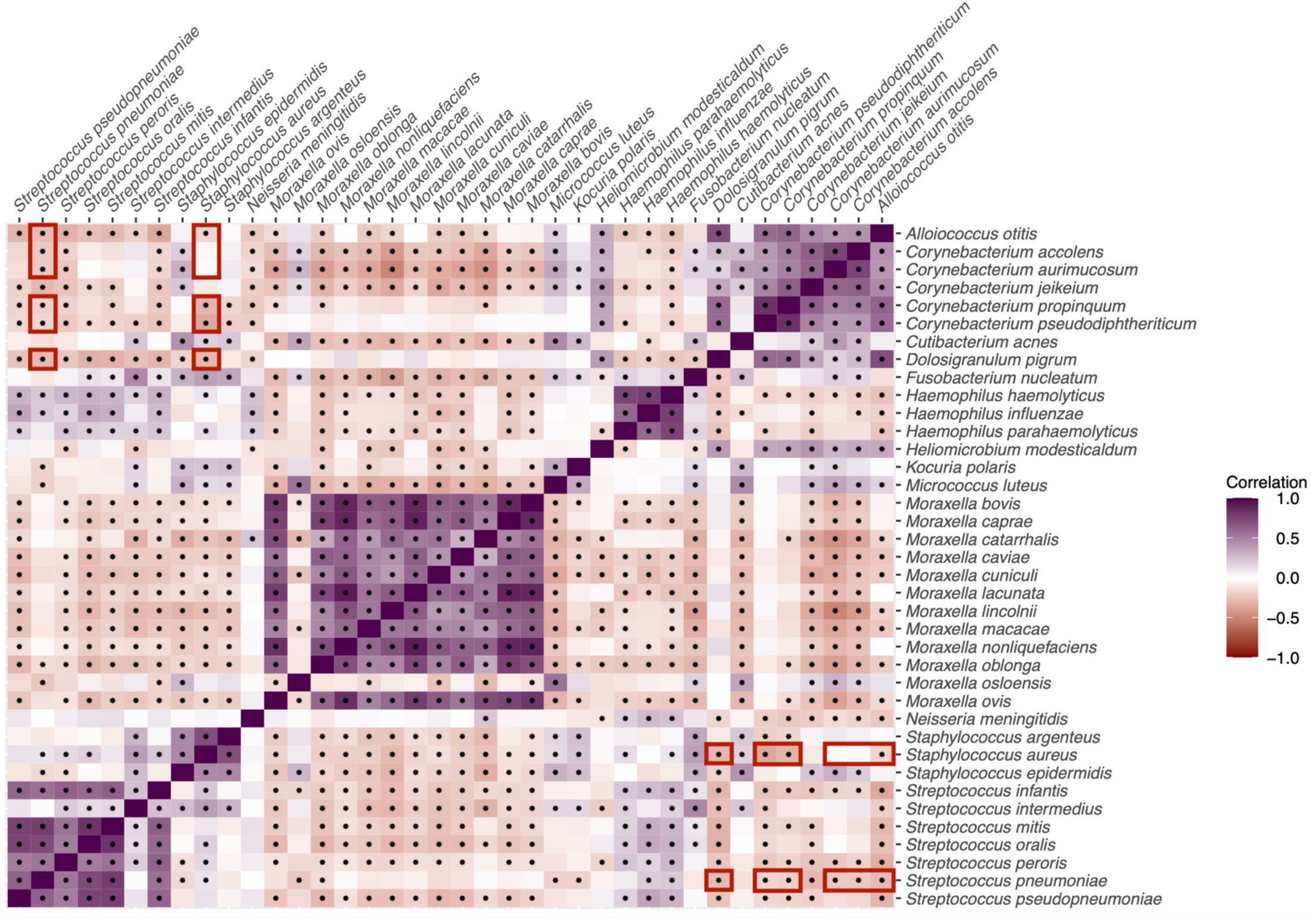
Interspecies correlations in the nasopharyngeal microbiomes of children in Botswana. Sparse Correlations for Compositional Data (SparCC) was used to generate interspecies correlations among species with ≥50 counts across the dataset. Each cell represents the correlation between two species, and species are arranged alphabetically. Positive correlations are indicated with purple shading and negative correlations are indicated with orange shading. Associations that remained significant after false discovery rate correction using Benjamini-Hochberg correction are marked with an asterisk. Associations between pathogens of interest (*S. pneumoniae*, *S. aureus*) and species differentially abundant by HIV status or HIV-associated immunosuppression are highlighted with maroon boxes.

## DISCUSSION

In this study, we described nasopharyngeal microbiome composition of children in Botswana and identified associations between HIV infection and microbiome composition not attributable to other factors like breastfeeding practices. We found that differences in microbiome composition among CLWH were driven primarily by HIV-associated immunosuppression. Finally, we demonstrated negative correlations between *Corynebacterium* and *Dolosigranulum* species that were less abundant in CLWH and the pathogens *S. pneumoniae* and *S. aureus*.

HIV infection is associated with a reduction of potentially protective bacterial species and an increase of potential pathogens in the nasopharynx. For example, CLWH with pneumonia in Botswana exhibited a near-complete absence of *D. pigrum* in nasopharyngeal samples.^10^ Additionally, other studies identified a higher prevalence of nasopharyngeal colonization with *S. pneumoniae, S. aureus,* and *H. influenzae* among CLWH.^5,6^ However, these studies were limited by enrollment of children with pneumonia and the use of culture-based identification methods. Importantly, we identified an association between HIV infection and nasopharyngeal microbiome composition among children without pneumonia in Botswana using shotgun metagenomic sequencing and both supervised and unsupervised clustering methods. While breastfeeding practices varied by HIV status in our study and breastfeeding affects nasopharyngeal microbiome composition in infancy, the differences in nasopharyngeal microbiome composition by HIV status persisted in a sub-analysis limited only to children with a history of breastfeeding. This suggests that differences in nasopharyngeal microbiome composition observed by HIV status among children in our study are not attributable to differences in breastfeeding practices. CLWH in our cohort also had a lower relative abundance of *C. pseudodiphtheriticum* compared to HUU children and a lower relative abundance of *C. accolens* compared to HEU children. Lastly, though we only had sequencing data for nine serodiscordant sibling pairs, the higher relative abundances of *C. propinquum*, *C. pseudodiphtheriticum,* and *D. pigrum* observed in the HEU siblings of CLWH and the higher abundance of pneumococcus in the CLWH suggest that differences in environmental exposures are unlikely to account for the nasopharyngeal microbiome alterations observed among CLWH. As *Corynebacterium* and *Dolosigranulum* species are associated with protection from respiratory infections in children, their lower relative abundance in CLWH may play a role in the increased incidence of pneumonia seen in this population.^24^

*D. pigrum* and multiple *Corynebacterium* species are common inhabitants of the upper respiratory tract and can influence colonization resistance to bacterial pathogens. *C. propinquum* inhibits growth of pneumococcus and some staphylococcal species *in vitro*, while *C. pseudodiphtheriticum* increases resistance to pneumococcal infection in mouse models and was successfully used for *S. aureus* eradication via nasal instillation in adults.^25–28^ *D. pigrum* also inhibits growth of *S. aureus*, and cocultivation of *C. pseudodiphtheriticum* and *D. pigrum* inhibits pneumococcal growth.^29^ Pneumococcus and *S. aureus* are responsible for significant childhood morbidity and mortality, and their ability to cause invasive disease is predicated on successful nasopharyngeal colonization.^1–3^ Among children in our study, the relative abundances of *C. propinquum*, *C. pseudodiphtheriticum* and *D. pigrum* were negatively correlated with the relative abundances of pneumococcus and *S. aureus*.

Within the context of the existing literature, our findings suggest that the lower nasopharyngeal abundance of *C. pseudodiphtheriticum* in CLWH and the lower relative abundances of *C. propinquum* and *C. pseudodiphtheriticum* in children with HIV-associated immunosuppression may contribute to the increased risk of invasive pneumococcal and *S. aureus* infections in this population.

Several factors likely contribute to the alterations of the nasopharyngeal microbiome observed among altered immunity and several prior studies suggest that HIV-associated immunosuppression influences microbiome composition. Among people with HIV, CD4+ cell counts are positively correlated with the relative abundances of *Streptococcus* and *Lactobacillus* in the salivary microbiome, while CD4+ cell counts <200 are associated with reduced alpha diversity and higher relative abundances of genera within the family *Enterobacteriaceae* in the gut microbiome.^30,31^ Similarly, low CD4+% was associated with alterations in nasopharyngeal microbiome composition in our cohort using both supervised and unsupervised clustering methods, and CLWH with normal CD4+% did not exhibit significant compositional differences compared to HEU and HUU children. Second, medications like ART may impact nasopharyngeal microbiome composition, as they are associated with alterations of the gut microbiome in CLWH.^8,32^ CLWH are also frequently exposed to antibiotics for both prophylaxis against opportunistic pathogens and treatment of acute infections, which can affect nasopharyngeal microbiome composition.^33^ In our cohort, receipt of TMP-SMX prophylaxis was associated with a higher relative abundance of *S. aureus*. Lastly, while associations exist between HIV RNA measurements and composition of other microbial communities, viral suppression was not consistently associated with nasopharyngeal microbiome composition in our cohort.^30^ Transient elevations in HIV RNA measurements can occur in children on ART and this may have been responsible for the detectable HIV RNA measurements among the children in our study, though the cross-sectional study design precludes us from assessing the duration of HIV viremia.^34^

Our study has several limitations. First, this study was cross-sectional, and we were thus unable to determine if the differences we observed in nasopharyngeal microbiome composition were associated with increased risk of subsequent respiratory infections in our study population. Due to the pediatric HIV clinical infrastructure in Botswana, we enrolled 43 (98%) CLWH from pediatric HIV subspecialty clinics, which limits our ability to assess for differences in microbiome composition by enrollment site. Of note, all enrollment sites were located within a 30-mile geographical radius. Additionally, the potential mode, inhaled medication use, and the impact of the COVID-19 pandemic on access to healthcare. Enrollment in this study was restricted to children without pneumonia or asthma in southeastern Botswana, and our results may not be generalizable to children with pneumonia or chronic pulmonary disease or to children in other settings, including other low- and middle-income countries where ART is not universally available and environmental and sociodemographic factors may differ. The use of shotgun metagenomic sequencing to characterize a low biomass microbial community carries the risk of not identifying all present species due to insufficient depth of coverage. However, we addressed this concern by processing our samples using a customized Kaiju pipeline modified for use with upper respiratory specimens. Furthermore, while the sequencing depth of our nasopharyngeal specimens was generally lower than datasets generated from samples from other body sites, it is quite robust when compared to other publicly available nasopharyngeal microbiome datasets.^35^ Our small sample size also limited our ability to adjust for potential confounders in analyses restricted to CLWH and sibling pairs; thus, these analyses should be considered exploratory. Lastly, we assessed differences in microbiome composition using several different statistical approaches, which if viewed independently carry a risk of overinterpretation.

In conclusion, HIV infection and HIV-associated immunosuppression were associated with alterations of the nasopharyngeal microbiomes of children in Botswana, including lower relative abundances of several putatively beneficial species whose abundances were negatively correlated with the abundances of the pathogens pneumococcus and *S. aureus.* Given the high burden of pneumonia among CLWH, there is an urgent need to develop strategies to modify the upper respiratory microbiome for the reduction or exclusion of potential pathogens and the prevention of respiratory infections in this population.

## ACKNOWLEDGMENTS

We thank Copan Italia (Brescia, Italy) for the donation of the MSwab media and flocked swabs used in the collection of nasopharyngeal specimens and the Duke University School of Medicine for the use of the Microbiome Core Facility and the Sequencing and Genomics Technologies Core Facility, which performed the DNA extractions and metagenomic sequencing for this research. Additionally, we thank the Botswana Ministry of Health, Greater Gaborone District Health Management Team, and clinic matrons for their support. Lastly, we offer our sincere gratitude to the children and families who participated in this research.

## DATA AVAILABILITY

All microbial genetic sequences generated as part of this study were uploaded to the National Center for Biotechnology Information and are available in the Sequence Read Archive under the BioProject Accession Number PRJNA1049442. Deidentified participant data can be made available via a shared GitHub repository after investigator approval of a proposal, which should be sent to the corresponding author. The R code used for analysis is available for review at https://github.com/swetamp/HIV_mb.

## ALT Text

Figure 1. Graphical representation of the screening and enrollment process, including numbers of children screened, excluded, and included in the study.

Figure 2. Principal component and stacked bar plots comparing microbiome composition by HIV status (among all children in the study) and immune suppression status (among children living with HIV).

Figure 3. Principal components plot and stacked bar plot displaying the five microbiome profiles generated by unsupervised clustering among all children in the study.

Figure 4. Plots of species in the nasopharyngeal microbiomes of Batswana children that were differentially abundant by sociodemographic factors, clinical factors, or environmental exposures with relative effect sizes and standard errors displayed.

Figure 5. Heat map displaying the strength of correlations between the most abundant species identified in the nasopharyngeal microbiomes of Batswana children.

