## Supplemental material for "Alterations of the upper respiratory microbiome among children living with HIV infection in Botswana"

Table of Contents | Page

| Table of Contents | Page |
|---|---|
| Statistical Appendix | 1 |
| Table S1. Species identified as probable sequencing contaminants using *decontam*, sorted by phylum. | 3 |
| Table S2. Most abundant phyla and genera isolated from study participant samples. | 5 |
| Table S3. Investigation of factors associated with alpha diversity measures using multivariable linear regression. | 6 |
| Table S4. Factors associated with altered nasopharyngeal microbiome composition using PERMANOVA. | 7 |
| Table S5. Characteristics of the study population by nasopharyngeal microbiome profile. | 8 |
| Table S6. Factors associated with microbiome profiles using multinomial logistic regression. | 9 |
| Table S7. Differentially abundant species by environmental and child characteristics identified using generalized linear models in MaAsLin2 and HUU as reference for HIV covariable. | 10 |
| Table S8. Differentially abundant species by environmental and child characteristics identified using generalized linear models in MaAsLin2 and HEU as reference for HIV covariable. | 11 |
| Table S9. Differentially abundant species by environmental and child characteristics among breastfed children identified using generalized linear models in MaAsLin2 and HEU as reference for HIV covariable. | 12 |
| Table S10. Baseline characteristics of sibling pairs (N=18). | 13 |
| Figure S1. Rarefaction curves used to determine pruning threshold for subsequent data analyses. | 14 |
| Figure S2. Alpha diversity of the nasopharyngeal microbiome among children in Botswana. | 15 |
| Figure S3. Composition of the nasopharyngeal microbiome by HIV status among children with a history of breastfeeding in Botswana. | 16 |
| Figure S4. Composition of the nasopharyngeal microbiome among sibling pairs. | 17 |
| Figure S5. Associations between HIV-associated factors and the composition of the microbiome among children in Botswana. | 18 |
| Figure S6. Composition of the nasopharyngeal microbiome by HIV immune status among children in Botswana. | 19 |
| Figure S7. Co-occurrence plot demonstrating associations between bacterial species in the nasopharyngeal microbiome. | 20 |
| Supplemental References | 21 |

### Statistical Appendix

We compared characteristics of CLWH, HEU children, and HUU children using Chi-squared or Fisher's exact tests for categorical variables and Kruskal-Wallis tests for continuous variables.

The primary outcome of interest in our analysis was differences in nasopharyngeal microbiome composition by HIV status. We evaluated differences in nasopharyngeal microbiome composition by assessing for 1) alpha, or within-sample diversity, 2) community-level or overall differences among sample groups, 3) microbiome profile differences among sample groups, and 4) differentially abundant taxa among sample groups.

**A.** To compare alpha diversity of samples by HIV status, we used the *phyloseq* R package version 1·38 to generate Shannon and Chao1 indices. We used multivariable linear regression to evaluate associations between HIV status and the Shannon index and log-transformed Chao1 richness of the microbiome. Analyses included our prespecified covariables of interest: season, age, household use of wood for cooking or heating, receipt of antibiotics within the preceding three months, and upper respiratory infection symptoms (nasal congestion, nasal discharge, or cough) in the preceding one month.

**B.** To compare the overall makeup of nasopharyngeal communities by HIV status, we first used the *microbiome* R package version 1·16 to generate centered log-ratio (CLR)-transformed sample counts to evaluate between-sample compositional differences. Employing CLR transformation addresses differences in sequencing depth between samples.[8] We generated PCoA plots of the transformed data grouped by HIV status using Euclidean distances. We then utilized permutational analysis of variance (PERMANOVA) with 1000 permutations to evaluate if the centroids of clusters of samples grouped by HIV status differed using Euclidean distances calculated from centered log-ratio (CLR)-transformed sample counts.[8] A separate PERMANOVA was run for each covariable of interest and each model was adjusted for age. All covariables that were significantly associated with overall microbiome composition were included in a single PERMANOVA to estimate the percentage of variance in microbiome composition explained by each.

**C.** To increase the rigor of our analyses that clustered microbiota by clinical variables, we also performed unsupervised clustering of taxonomic data to generate microbiome profiles and then evaluated if microbiome profiles were associated with HIV status. To generate nasopharyngeal microbiome profiles, we used k-medoids clustering followed by the Calinski-Harabasz index to identify the optimal number of clusters.[9,10] The use of k-medoids clustering, also known as partitioning around medoids (PAM), is a widely used and robust unsupervised clustering method used to identify naturally occurring clusters within microbiome data.[17,18] The Calinski-Harabasz index is a variance ratio criterion used to determine the optimal number of clusters generated from a clustering analysis and is often used in conjunction with PAM in microbiome analyses.[19,20] We examined univariable associations between clinical variables and nasopharyngeal microbiome profile using Chi-squared or Fisher's exact tests for categorical variables and Kruskal-Wallis tests for continuous variables. We examined multivariable associations between our covariables of interest and microbiome profile using multinomial logistic regression.

**D.** To identify differentially abundant species by HIV status, we filtered taxa with fewer than 50 counts across the dataset and those that were present in <20% of samples and fit multivariable general linear models using MaAsLin 2·0, which allowed for the inclusion of our prespecified covariables and correction for multiple testing.[11] We used a q value cutoff of 0·2 and adjusted for multiple testing using the Benjamini-Hochberg procedure. We utilized MaAsLin 2·0 instead of another method for identifying differential abundant species like DESeq2 due to concerns regarding unacceptably high false discovery rates. Two recent studies comparing several differential abundance analytic methods found improved performance with MaAsLin2 over DESeq2 due to the inability of the latter to adequately control the false discovery rate.[12,13] As MaAsLin only supports pairwise comparisons, we ran one model with HUU as the reference for the HIV status covariable to identify species that were differentially abundant in CLWH or HEU children compared to HUU children, and we ran a second model with HEU as the reference for the HIV status covariable to identify species that were differentially abundant in CLWH compared to HEU children. Both models utilized the aforementioned parameters and covariables of interest.

**E.** We also conducted multiple sub-analyses of our data to better understand the association we identified between HIV and microbiome composition:

a. Sensitivity analysis of microbiome composition comparison by HIV status, restricted to children with a history of exclusive breastfeeding. We used the same statistical methods as in analyses of the entire cohort.

b. Comparison of microbiome composition between CLWH and their HEU siblings (n=18). We compared microbiome composition among CLWH and their HEU siblings under five years of age to more precisely identify associations between HIV infection and microbiome composition, given that environmental exposures among these sibling pairs were expected to be similar.[14-16] We used paired PERMANOVA testing to compare microbiome composition within sibling pairs by covariables of interest expected to differ between sibling pairs (age, season, and upper respiratory infection symptoms in the preceding one month). Paired PERMANOVA allows for multilevel pairwise comparisons and adjusts for multiple testing using the Benjamini Hochberg procedure.[17] We did not compare composition by antibiotic exposure because only three children had received antibiotics in the preceding three months. Due to the small sample size (n=18 children grouped into nine pairs), we ran only univariable paired PERMANOVA models and did not adjust for potential confounders. Due to the small sample size and paired data structure, we used Wilcoxon signed-rank tests to compare the relative abundances of the ten most common bacterial species within sibling pairs.

c. Identification of differentially abundant taxa among CLWH (n=44). We compared overall microbiome composition by CD4+% classification (normal vs low) using PERMANOVA. After filtering the dataset as described above, we used the linear discriminant effect size (LEfSe) method to identify differentially abundant species by CD4+ cell percentage, viral load, use of trimethoprim-sulfamethoxazole (TMP-SMX) prophylaxis, and antibiotic treatment in the preceding three months.[18] We chose LEfSe for these analyses over multivariable general linear models as our small sample size limited our ability to adjust for confounders and because LEfSe is widely used for differential abundance analyses, including those involving the nasopharyngeal microbiome.[19,20] LEfSe utilizes a Kruskal-Wallis rank sum test to identify species that are differentially abundant among classes of interest (e.g., CD4+% classification) and then utilizes an linear discriminant analysis (LDA) model that incorporates bootstrapping over 30 cycles to determine the effect size associated with each differentially abundant species. Specifically, effect sizes are determined by taking the average of two values: 1) the difference between class means and 2) the difference between class means along the first linear discriminant axis, which takes into account the variability and discriminatory power of each differentially abundant species. This value is then scaled in the $[1, 10^6]$ interval and the LDA score is generated by taking the logarithm of the scaled value. LDA scores generated by the LEfSe method provide an estimation of the order of magnitude difference in the relative abundance of a species that can be attributed to the variable of interest (e.g., CD4+% classification or antibiotic exposure). LEfSe was validated using several datasets, including one containing only thirteen metagenomes, which supports the strength of this analytic approach with smaller sample sizes.[18] We used the *microbiomeMarker* R package version 1·0·2 to perform our LEfSe analyses using a Kruskal-Wallis cutoff of alpha = 0·01 and an LDA cutoff of 2.

d. Comparison of microbiome composition between immunosuppressed CLWH (n=11) and HEU and HUU children. We used PERMANOVA to compare microbiome composition among the three groups in this subanalysis. As one of our comparison groups contained only n=11 children, we used LEfSe to identify differentially abundant species using the parameters described in (c).

e. Comparison of microbiome composition between immunocompetent CLWH (n=31) and HEU and HUU children. To maintain consistency with our other subanalyses, we used PERMANOVA to compare microbiome composition among the three groups in this subanalysis and LEfSe to identify differentially abundant species using the parameters described in (c).

f. Comparison of unsupervised cluster membership among CLWH. We performed subgroup analyses of our unsupervised clusters restricted to CLWH to identify clusters associated with viral suppression and CD4+% classification. Due to the small sample size of children included in these subanalyses, we divided the five microbiome profiles into two groups and used Chi-squared or Fisher's exact tests to identify associations with viral suppression and CD4+% classification.

**F.** Lastly, we filtered taxa with fewer than 50 counts across the dataset and used the *SpiecEasi* R package version 1·1·2 to generate Sparse Correlations for Compositional Data (SparCC) with 1000 bootstrap replicates and adjustment for false discovery due to multiple testing using the Benjamini Hochberg correction. SparCC identifies pairwise correlations between specific bacterial species using log-transformed relative abundance data and an iterative process. We chose SparCC because it was developed to identify correlations among compositional data and adjusts for multiple testing.[21] We next generated a co-occurrence network from our SparCC results using Cytoscape version 3·10.[22]

**Table S1. Species identified as probable sequencing contaminants using *decontam*.**

| Kingdom | Phylum | Class | Order | Family | Genus | Species |
|---|---|---|---|---|---|---|
| Bacteria | Actinobacteria | Actinomycetia | Bifidobacteriales | Bifidobacteriaceae | *Bifidobacterium* | *Bifidobacterium subtile* |
| Bacteria | Actinobacteria | Actinomycetia | Frankiales | Frankiaceae | *Frankia* | *Frankia discariae* |
| Bacteria | Actinobacteria | Actinomycetia | Micrococcales | Micrococcaceae | *Arthrobacter* | *Arthrobacter sp. 31Y* |
| Bacteria | Actinobacteria | Actinomycetia | Pseudonocardiales | Pseudonocardiaceae | *Saccharothrix* | *Saccharothrix sp. NRRL B-16348* |
| Bacteria | Bacteroidetes | Bacteroidia | Bacteroidales | Prevotellaceae | *Prevotella* | *Prevotella sp. lc2012* |
| Bacteria | Bacteroidetes | Bacteroidia | Bacteroidales | Rikenellaceae | *Alistipes* | *Alistipes sp. ZOR0009* |
| Bacteria | Bacteroidetes | Cytophagia | Cytophagales | Cyclobacteriaceae | *Echinicola* | *Echinicola pacifica* |
| Bacteria | Bacteroidetes | Cytophagia | Cytophagales | Cyclobacteriaceae | *Cyclobacterium* | *Cyclobacterium lianum* |
| Bacteria | Bacteroidetes | Flavobacteriia | Flavobacteriales | Flavobacteriaceae | *Flavobacterium* | *Flavobacterium fontis* |
| Bacteria | Bacteroidetes | Flavobacteriia | Flavobacteriales | Flavobacteriaceae | *Flavobacterium* | *Flavobacterium psychrophilum* |
| Bacteria | Bacteroidetes | Flavobacteriia | Flavobacteriales | Flavobacteriaceae | *Flavobacterium* | *Flavobacterium phragmitis* |
| Bacteria | Bacteroidetes | Flavobacteriia | Flavobacteriales | Weeksellaceae | *Chryseobacterium* | *Chryseobacterium jejuense* |
| Bacteria | Bacteroidetes | Flavobacteriia | Flavobacteriales | Weeksellaceae | *Chryseobacterium* | *Chryseobacterium joostei* |
| Bacteria | Bacteroidetes | Sphingobacteriia | Sphingobacteriales | Sphingobacteriaceae | *Sphingobacterium* | *Sphingobacterium sp. 21* |
| Bacteria | Bacteroidetes | Sphingobacteriia | Sphingobacteriales | Sphingobacteriaceae | *Sphingobacterium* | *Sphingobacterium paucimobilis* |
| Bacteria | Candidatus Woesebacteria | NA | NA | NA | NA | *Candidatus Woesebacteria bacterium GW2011_GWF1_31_35* |
| Bacteria | Deinococcus-Thermus | Deinococci | Deinococcales | Deinococcaceae | *Deinococcus* | *Deinococcus phoenicis* |
| Bacteria | Deinococcus-Thermus | Deinococci | Thermales | Thermaceae | *Thermus* | *Thermus antranikianii* |
| Bacteria | Firmicutes | Bacilli | Bacillales | Bacillaceae | *Peribacillus* | *Peribacillus kribbensis* |
| Bacteria | Firmicutes | Bacilli | Bacillales | Bacillaceae | *Alteribacillus* | *Alteribacillus persepolensis* |
| Bacteria | Firmicutes | Bacilli | Bacillales | Bacillaceae | *Caldibacillus* | *Caldibacillus thermoamylovorans* |
| Bacteria | Firmicutes | Bacilli | Bacillales | Bacillaceae | *Schinkia* | *Schinkia azotoformans* |
| Bacteria | Firmicutes | Bacilli | Bacillales | Paenibacillaceae | *Paenibacillus* | *Paenibacillus oryzae* |
| Bacteria | Firmicutes | Bacilli | Lactobacillales | Lactobacillaceae | *Lactobacillus* | *Lactobacillus johnsonii* |
| Bacteria | Firmicutes | Bacilli | Lactobacillales | Lactobacillaceae | *Lactiplantibacillus* | *Lactiplantibacillus pentosus* |
| Bacteria | Firmicutes | Clostridia | Eubacteriales | NA | NA | *Clostridiales bacterium oral taxon 876* |
| Bacteria | Fusobacteria | Fusobacteriia | Fusobacteriales | Leptotrichiaceae | *Streptobacillus* | *Streptobacillus ratti* |
| Bacteria | Fusobacteria | Fusobacteriia | Fusobacteriales | Leptotrichiaceae | *Streptobacillus* | *Streptobacillus moniliformis* |
| Bacteria | Proteobacteria | Alphaproteobacteria | Hyphomicrobiales | NA | NA | *Rhizobiales bacterium Ga0077525* |
| Bacteria | Proteobacteria | Alphaproteobacteria | Hyphomicrobiales | Rhizobiaceae | *Rhizobium* | *Rhizobium tibeticum* |
| Bacteria | Proteobacteria | Alphaproteobacteria | Rhodobacterales | NA | NA | *Rhodobacterales bacterium RIFCSPHIGHO2_02_FULL_62_130* |
| Bacteria | Proteobacteria | Alphaproteobacteria | Rhodobacterales | Rhodobacteraceae | *Pseudorhodobacter* | *Pseudorhodobacter aquimaris* |
| Bacteria | Proteobacteria | Alphaproteobacteria | Rhodobacterales | Rhodobacteraceae | *Puniceibacterium* | *Puniceibacterium sp. IMCC21224* |
| Bacteria | Proteobacteria | Alphaproteobacteria | Rhodospirillales | Acetobacteraceae | *Kozakia* | *Kozakia baliensis* |
| Bacteria | Proteobacteria | Alphaproteobacteria | Sphingomonadales | Sphingomonadaceae | *Novosphingobium* | *Novosphingobium resinovorum* |
| Bacteria | Proteobacteria | Alphaproteobacteria | Sphingomonadales | Sphingomonadaceae | *Sphingomonas* | *Sphingomonas sp. Leaf343* |
| Bacteria | Proteobacteria | Betaproteobacteria | NA | NA | *Candidatus Accumulibacter* | *Candidatus Accumulibacter sp. BA-93* |
| Bacteria | Proteobacteria | Betaproteobacteria | Burkholderiales | Alcaligenaceae | *Bordetella* | *Bordetella trematum* |
| Bacteria | Proteobacteria | Betaproteobacteria | Burkholderiales | Comamonadaceae | *Acidovorax* | *Acidovorax sp. NO-1* |
| Bacteria | Proteobacteria | Betaproteobacteria | Burkholderiales | Comamonadaceae | *Hydrogenophaga* | *Hydrogenophaga sp. IBVHS1* |
| Bacteria | Proteobacteria | Betaproteobacteria | Burkholderiales | Comamonadaceae | *Hydrogenophaga* | *Hydrogenophaga sp. A37* |
| Bacteria | Proteobacteria | Betaproteobacteria | Burkholderiales | Comamonadaceae | *Hydrogenophaga* | *Hydrogenophaga sp. IBVHS2* |
| Bacteria | Proteobacteria | Betaproteobacteria | Burkholderiales | Comamonadaceae | *Pseudacidovorax* | *Pseudacidovorax sp. RU35E* |
| Bacteria | Proteobacteria | Betaproteobacteria | Burkholderiales | Comamonadaceae | *Verminephrobacter* | *Verminephrobacter eiseniae* |
| Bacteria | Proteobacteria | Betaproteobacteria | Burkholderiales | Oxalobacteraceae | *Janthinobacterium* | *Janthinobacterium agaricidamnosum* |
| Bacteria | Proteobacteria | Gammaproteobacteria | Alteromonadales | Idiomarinaceae | *Idiomarina* | *Idiomarina sp. 5.13* |
| Bacteria | Proteobacteria | Gammaproteobacteria | Enterobacterales | Yersiniaceae | *Serratia* | *Serratia sp. Ag1* |
| Bacteria | Proteobacteria | Gammaproteobacteria | Enterobacterales | Erwiniaceae | *Erwinia* | *Erwinia tracheiphila* |

| | | | | | | |
|---|---|---|---|---|---|---|
| Bacteria | Proteobacteria | Gammaproteobacteria | NA | NA | NA | *Gammaproteobacteria bacterium RIFCSPHIGHO2_12_FULL_38_11* |
| Bacteria | Proteobacteria | Gammaproteobacteria | Pseudomonadales | Pseudomonadaceae | *Pseudomonas* | *Pseudomonas sp. URHB0015* |
| Bacteria | Proteobacteria | Gammaproteobacteria | Xanthomonadales | Rhodanobacteraceae | *Dokdonella* | *Dokdonella koreensis* |
| Bacteria | Spirochaetes | Spirochaetia | Spirochaetales | Treponemataceae | *Treponema* | *Treponema bryantii* |
| Bacteria | Thermotogae | Thermotogae | Thermotogales | Fervidobacteriaceae | *Fervidobacterium* | *Fervidobacterium nodosum* |

**Table S2. Most abundant phyla and genera detected in samples**

| Phyla and genera | | Relative abundance | |
|---|---|---|---|
| Proteobacteria | | 0·37 | |
| | *Moraxella* | | 0·30 |
| | *Haemophilus* | | 0·04 |
| | *Actinobacillus* | | 0·004 |
| | Other genera | | 0·026 |
| Firmicutes | | 0·32 | |
| | *Dolosigranulum* | | 0·20 |
| | *Streptococcus* | | 0·07 |
| | *Staphylococcus* | | 0·03 |
| | Other genera | | 0·02 |
| Actinobacteria | | 0·30 | |
| | *Corynebacterium* | | 0·26 |
| | *Micrococcus* | | 0·01 |
| | *Cutibacterium* | | 0·005 |
| | *Kocuria* | | 0·004 |
| | Other genera | | 0·021 |
| Other phyla | | 0·01 | |

**Table S3. Investigation of factors associated with alpha diversity measures using multivariable linear regression**

| Factor | Estimate | 95% CI | p |
|---|---|---|---|
| ***Shannon index*** | | | |
| HIV infection (CLWH) | 0.024 | -0.230, 0.277 | 0.85 |
| HIV exposure (HEU children) | 0.153 | -0.095, 0.400 | 0.22 |
| Wood smoke exposure | 0.044 | -0.172, 0.260 | 0.69 |
| Age | -0.0003 | -0.007, 0.006 | 0.92 |
| Recent URI symptoms | -0.118 | -0.332, 0.097 | 0.28 |
| Antibiotic exposure in prior 3 months | 0.067 | -0.213, 0.347 | 0.64 |
| Rainy season (November to March) | 0.055 | -0.161, 0.270 | 0.62 |
| ***log Chao1*** | | | |
| HIV infection (CLWH) | -0.137 | -0.571, 0.297 | 0.53 |
| HIV exposure (HEU children) | 0.249 | -0.174, 0.672 | 0.25 |
| Wood smoke exposure | -0.012 | -0.382, 0.357 | 0.95 |
| Age | -0.005 | -0.016, 0.006 | 0.34 |
| Recent URI symptoms | -0.071 | -0.438, 0.297 | 0.70 |
| Antibiotic exposure in prior 3 months | -0.040 | -0.519, 0.438 | 0.87 |
| Rainy season (November to March) | -0.136 | -0.505, 0.232 | 0.47 |

CI, confidence interval; URI, upper respiratory infection

**Table S4. Factors associated with altered nasopharyngeal microbiome composition using PERMANOVA**

| **Characteristic** | **$R^2$** | **p** |
|---|---|---|
| HIV infection | 0·019 | 0·046 |
| Season | 0·016 | 0·0040 |
| Recent URI symptoms | 0·013 | 0·0080 |
| Age | 0·011 | 0·037 |

URI, upper respiratory infection

### Table S5. Characteristics of the study population by nasopharyngeal microbiome profile

| | Microbiome profile | | | | | |
|---|---|---|---|---|---|---|
| | CPD: *Corynebacterium pseudodiphtheriticum*/ *Dolosigranulum pigrum* dominant (n=44) | BIO: Biodiverse (n=27) | MCA: *Moraxella catarrhalis* dominant (n=16) | MNO: *M. nonliquefaciens* dominant (n=29) | CPR: *C. propinquum* dominant (n=27) | p* |
| | N (%) | N (%) | N (%) | N (%) | N (%) | |
| **Demographics** | | | | | | |
| Child age in months, median (IQR) | 36 (19, 49) | 30 (14, 46) | 18 (14, 26) | 36 (28, 49) | 37 (20, 53) | 0·042 |
| Female sex | 24 (55) | 14 (52) | 7 (44) | 16 (55) | 13 (48) | 0·94 |
| HIV status | | | | | | 0·17 |
| Children living with HIV | 12 (27) | 14 (52) | 4 (25) | 9 (31) | 5 (19) | |
| HIV-exposed, uninfected | 14 (32) | 9 (33) | 5 (31) | 8 (28) | 13(48) | |
| HIV-unexposed, uninfected | 18 (41) | 4 (15) | 7 (44) | 12 (41) | 9 (33) | |
| Maternal age, y, median (IQR) | 28 (24, 35) | 29 (25, 35) | 26 (25, 31) | 30 (27, 35) | 30 (25, 38) | 0·67 |
| Rainy season (November to March) | 31 (70) | 18 (67) | 10 (63) | 10 (34) | 21 (78) | 0·0078 |
| **Socioeconomic factors** | | | | | | |
| Maternal education level (N=142) | | | | | | 0·59 |
| None or primary | 3 (7) | 5 (19) | 0 (0) | 5 (18) | 2 (7) | |
| Secondary | 34 (77) | 20 (74) | 14 (87) | 20 (71) | 22 (82) | |
| Tertiary | 7 (16) | 2 (7) | 2 (13) | 3 (11) | 3 (11) | |
| Electricity in home | 33 (75) | 23 (85) | 9 (56) | 19 (66) | 22 (81) | 0·19 |
| Use of wood for cooking or heating | 21 (48) | 21 (78) | 10 (63) | 20 (69) | 15 (56) | 0·11 |
| Number of household members, median (IQR) | 4 (3,6) | 6 (4,8) | 4 (4,5) | 4 (3,5) | 4 (3,5) | 0·0022 |
| **Clinical factors** | | | | | | |
| Antibiotic exposure in prior 3 months | 4 (9) | 8 (30) | 4 (25) | 6 (21) | 2 (7) | 0·087 |
| URI symptoms in prior 1 month | 13 (30) | 15 (56) | 11 (69) | 10 (34) | 10 (37) | 0·033 |
| Received 3 doses of PCV-13 (N=141) | 40 (93) | 22 (85) | 14 (88) | 27 (93) | 24 (89) | 0·77 |
| Received 3 doses of Hib vaccine (N=141) | 39 (91) | 24 (92) | 15 (94) | 27 (93) | 25 (93) | 1·0 |
| Ever breastfed | 26 (59) | 11 (41) | 10 (63) | 21 (72) | 16 (59) | 0·20 |
| Weight-for-age z-scores, median (IQR) | -0·13 (-0·89, 0·18) | -0·75 (-1·45, -0·14) | -1·19 (-1·71, 0·24) | -0·27 (-1·.07, 0·01) | 0·05 (-0·67, 0·36) | 0·060 |
| Height-for-age z-scores, median (IQR) | -0·08 (-0·90, 0·64) | -0·82 (-1·50, 0·01) | -0·74 (-1·91, -0·05) | -0·60 (-1·29, 0·56) | -0·13 (-1·11, 0·69) | 0·15 |

IQR, interquartile range; URI, upper respiratory infection; PCV-13, 13-valent pneumococcal conjugate vaccine; Hib, *H. influenzae* type B
**p* values were estimated using Chi-square or Fisher's exact tests for categorical variables and Kruskal-Wallis test for continuous variables

**Table S6. Factors associated with microbiome profiles using multinomial logistic regression**

| Microbiome profile | Factor | Relative risk ratio (95% CI) | p |
|---|---|---|---|
| BIO: Biodiverse | HIV infection | 5·54 (1·37, 22·33) | 0·016 |
| BIO: Biodiverse | Wood smoke exposure | 3·34 (1·05, 10·56) | 0·040 |
| MCA: *Moraxella catarrhalis* dominant | Age | 0·94 (0·90, 0·98) | 0·0078 |
| MCA: *Moraxella catarrhalis* dominant | Recent URI symptoms | 4·54 (1·24, 16·67) | 0·022 |
| MNO: *M. nonliquefaciens* dominant | Rainy season (November to March) | 0·23 (0·08, 0·65) | 0·0056 |

CI, confidence interval; URI, upper respiratory infection

**Table S7. Differentially abundant species by environmental and child characteristics identified using generalized linear models in MaAsLin2 and HUU as reference for HIV covariable**

| | Effect estimate | Standard error | p value | q value* |
|---|---|---|---|---|
| **Rainy season** | | | | |
| *Moraxella ovis* | -2·31 | 0·559 | <0·0001 | 0·0031 |
| *Moraxella oblonga* | -1·95 | 0·557 | 0·0006 | 0·018 |
| *Moraxella nonliquefaciens* | -3·48 | 0·940 | 0·0003 | 0·011 |
| *Moraxella macacae* | -1·16 | 0·409 | 0·0053 | 0·087 |
| *Moraxella lincolnii* | -1·94 | 0·596 | 0·0014 | 0·031 |
| *Moraxella lacunata* | -3·53 | 0·829 | <0·0001 | 0·0029 |
| *Moraxella cuniculi* | -1·96 | 0·513 | 0·0002 | 0·0081 |
| *Moraxella caviae* | -2·37 | 0·563 | <0·0001 | 0·0029 |
| *Moraxella catarrhalis* | -1·72 | 0·667 | 0·011 | 0·159 |
| *Moraxella caprae* | -2·64 | 0·572 | <0·0001 | 0·0021 |
| *Moraxella bovis* | -2·49 | 0·580 | <0·0001 | 0·0029 |
| *Kocuria polaris* | -1·42 | 0·495 | 0·0048 | 0·085 |
| *Micrococcus luteus* | 1·77 | 0·558 | 0·0019 | 0·039 |
| **HIV infection** | | | | |
| *Corynebacterium pseudodiphtheriticum* | -2·22 | 0·848 | 0·010 | 0·15 |
| **Recent URI symptoms** | | | | |
| *Moraxella cuniculi* | 1·27 | 0·511 | 0·014 | 0·20 |
| **Age** | | | | |
| *Streptococcus peroris* | -0·81 | 0·223 | 0·0004 | 0·012 |
| *Streptococcus infantis* | -0·65 | 0·223 | 0·0041 | 0·077 |
| *Micrococcus luteus* | 0·91 | 0·270 | 0·0010 | 0·024 |

URI, upper respiratory infection; HUU, HIV-unexposed, uninfected.

*q values refer to adjusted significance using Benjamini-Hochberg Procedure to account for the false discovery rate

**Table S8. Differentially abundant species by environmental and child characteristics identified using generalized linear models in MaAsLin2 and HEU as reference for HIV covariable**

| | Effect estimate | Standard error | p value | q value* |
|---|---|---|---|---|
| **Rainy season** | | | | |
| *Moraxella ovis* | -2·31 | 0·559 | <0·0001 | 0·0031 |
| *Moraxella oblonga* | -1·95 | 0·557 | 0·0006 | 0·016 |
| *Moraxella nonliquefaciens* | -3·48 | 0·940 | 0·0003 | 0·011 |
| *Moraxella macacae* | -1·16 | 0·409 | 0·0053 | 0·082 |
| *Moraxella lincolnii* | -1·94 | 0·596 | 0·0014 | 0·029 |
| *Moraxella lacunata* | -3·53 | 0·829 | <0·0001 | 0·0029 |
| *Moraxella cuniculi* | -1·96 | 0·513 | 0·0002 | 0·0081 |
| *Moraxella caviae* | -2·37 | 0·563 | <0·0001 | 0·0029 |
| *Moraxella catarrhalis* | -1·72 | 0·667 | 0·011 | 0·14 |
| *Moraxella caprae* | -2·64 | 0·572 | <0·0001 | 0·0021 |
| *Moraxella bovis* | -2·49 | 0·580 | <0·0001 | 0·0029 |
| *Kocuria polaris* | -1·42 | 0·495 | 0·0048 | 0·079 |
| *Moraxella osloensis* | 1·38 | 0·567 | 0·016 | 0·19 |
| *Micrococcus luteus* | 1·77 | 0·558 | 0·0019 | 0·036 |
| **HIV infection** | | | | |
| *Corynebacterium accolens* | -2·06 | 0·743 | 0·0063 | 0·091 |
| *Corynebacterium aurimucosum* | -2·07 | 0·588 | 0·0006 | 0·016 |
| *Micrococcus luteus* | -1·76 | 0·661 | 0·0085 | 0·12 |
| **Recent URI symptoms** | | | | |
| *Moraxella cuniculi* | 1·27 | 0·511 | 0·014 | 0·18 |
| *Haemophilus parahaemolyticus* | 1·20 | 0·500 | 0·018 | 0·19 |
| **Age** | | | | |
| *Streptococcus peroris* | -0·81 | 0·223 | 0·0004 | 0·012 |
| *Streptococcus infantis* | -0·65 | 0·223 | 0·0041 | 0·071 |
| *Micrococcus luteus* | 0·91 | 0·270 | 0·0010 | 0·022 |
| *Kocuria polaris* | 0·57 | 0·239 | 0·018 | 0·19 |

URI, upper respiratory infection; HEU, HIV-exposed, uninfected.

*q values refer to adjusted significance using Benjamini-Hochberg Procedure to account for the false discovery rate

**Table S9. Differentially abundant species by environmental and child characteristics among breastfed children identified using generalized linear models in MaAsLin2 and HEU as reference for HIV covariable**

| | Effect estimate | Standard error | p value | q value* |
|---|---|---|---|---|
| **Rainy season** | | | | |
| *Moraxella bovis* | -2·40 | 0·77 | 0·0026 | 0·087 |
| *Moraxella caprae* | -2·61 | 0·77 | 0·0011 | 0·079 |
| *Moraxella lacunata* | -3·46 | 1·12 | 0·0029 | 0·087 |
| *Moraxella nonliquefaciens* | -3·78 | 1·27 | 0·0040 | 0·095 |
| **HIV infection** | | | | |
| *Corynebacterium accolens* | -4·06 | 1·07 | 0·0003 | 0·030 |
| *Corynebacterium aurimucosum* | -3·10 | 0·94 | 0·0015 | 0·079 |
| *Micrococcus luteus* | -3·84 | 0·95 | 0·0001 | 0·027 |
| *Moraxella ovis* | 3·23 | 1·06 | 0·0032 | 0·087 |
| **Age** | | | | |
| *Micrococcus luteus* | 0·97 | 0·32 | 0·0030 | 0·087 |

HEU, HIV-exposed, uninfected.
*q values refer to adjusted significance using Benjamini-Hochberg Procedure to account for the false discovery rate

**Table S10. Baseline characteristics of sibling pairs (N=18)**

| | Overall (n=18) | Children with HIV (n=9) | HEU siblings (n=9) | p* |
|---|---|---|---|---|
| | N (%) | N (%) | N (%) | |
| **Demographics** | | | | |
| Child age in months, median (IQR) | 25 (9, 42) | 30 (19, 39) | 19 (7, 43) | 0·59 |
| Female sex | 11 (61) | 5 (56) | 6 (67) | 0·72 |
| Rainy season (November to March) | 9 (50) | 4 (44) | 5 (56) | 1·0 |
| **Clinical factors** | | | | |
| Antibiotic exposure in prior 3 months | 3 (17) | 1 (11) | 2 (22) | 0·077 |
| URI symptoms in prior 1 month | 11 (61) | 6 (67) | 5 (56) | 0·75 |
| Received 3 doses of PCV-13 (N=141) | 14 (78) | 7 (78) | 7 (78) | 0·85 |
| Received 3 doses of Hib vaccine (N=141) | 14 (78) | 7 (78) | 7 (78) | 0·85 |
| History of any breastfeeding | 8 (44) | 5 (56) | 3 (33) | 1·0 |
| Weight-for-age z-scores, median (IQR) | -0·47 (-1·49, 0·73) | -1·24 (-1·69, -0·70) | 0·58 (-0·23, 0·92) | 0·024 |
| Height-for-age z-scores, median (IQR) | -0·92 (-1·54, 0·07) | -1·22 (-1·90, -0·88) | -0·33 (-0·96, 0·81) | 0·033 |

HEU, HIV-exposed, uninfected; IQR, interquartile range; URI, upper respiratory infection; PCV-13, 13-valent pneumococcal conjugate vaccine; Hib, *H. influenzae* type B
*p values calculated using Wilcoxon signed-rank tests for continuous variables and McNemar's test for categorical variables.

**Figure S1. Rarefaction curves used to determine pruning threshold for subsequent data analyses.** The *phyloseq* package in R was used to subsample each specimen ten times at a range of sequencing depths to generate curves of Shannon and Simpson diversity indices. Sample-specific rarefaction curves are shown.

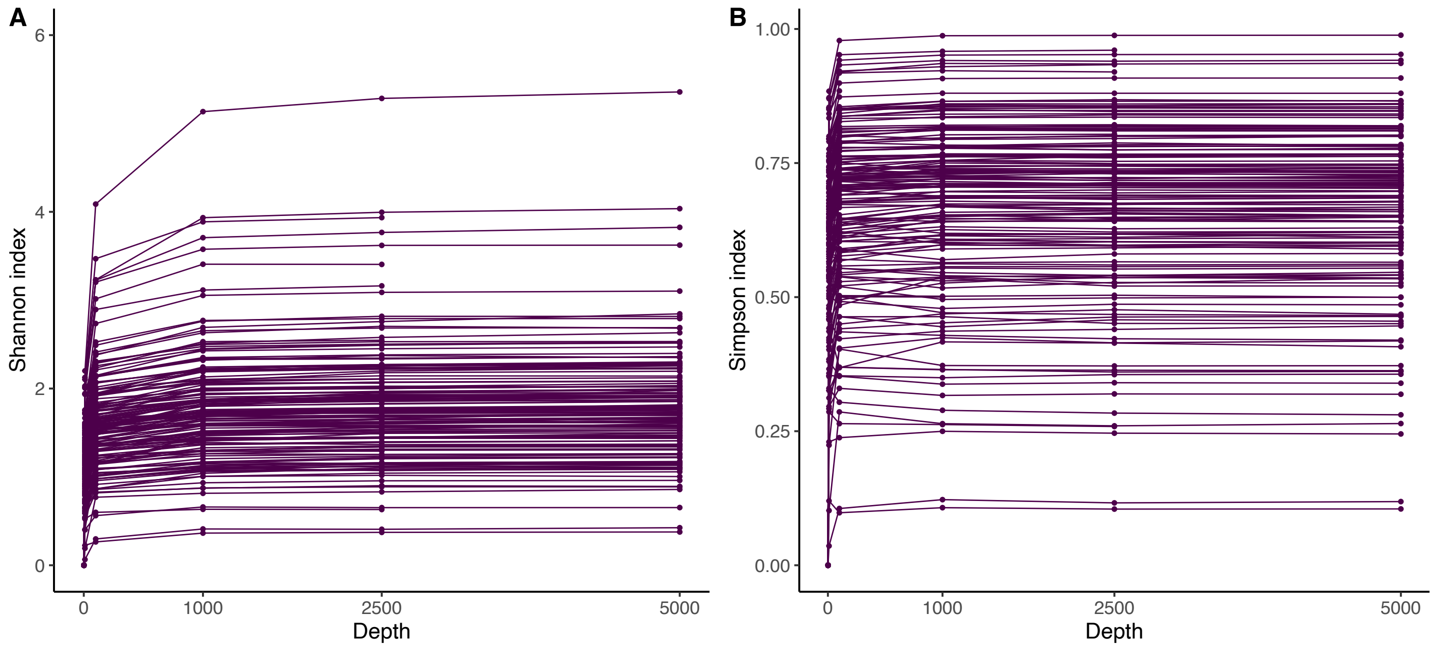

**Figure S2. Alpha diversity of the nasopharyngeal microbiome among children in Botswana.** Box plots display nasopharyngeal microbiome diversity, as measured by the Shannon index (A), and richness, as measured by the Chao1 richness (B), by HIV status. The center line of each box corresponds to the median value and the upper and lower bounds of each box correspond to the first and third quartiles. Whiskers represent values within 1.5x the interquartile range and outliers are represented as points. No significant differences in alpha diversity measures were observed by HIV status. CLWH, children living with HIV; HEU, HIV-exposed, uninfected; HUU, HIV-unexposed, uninfected.

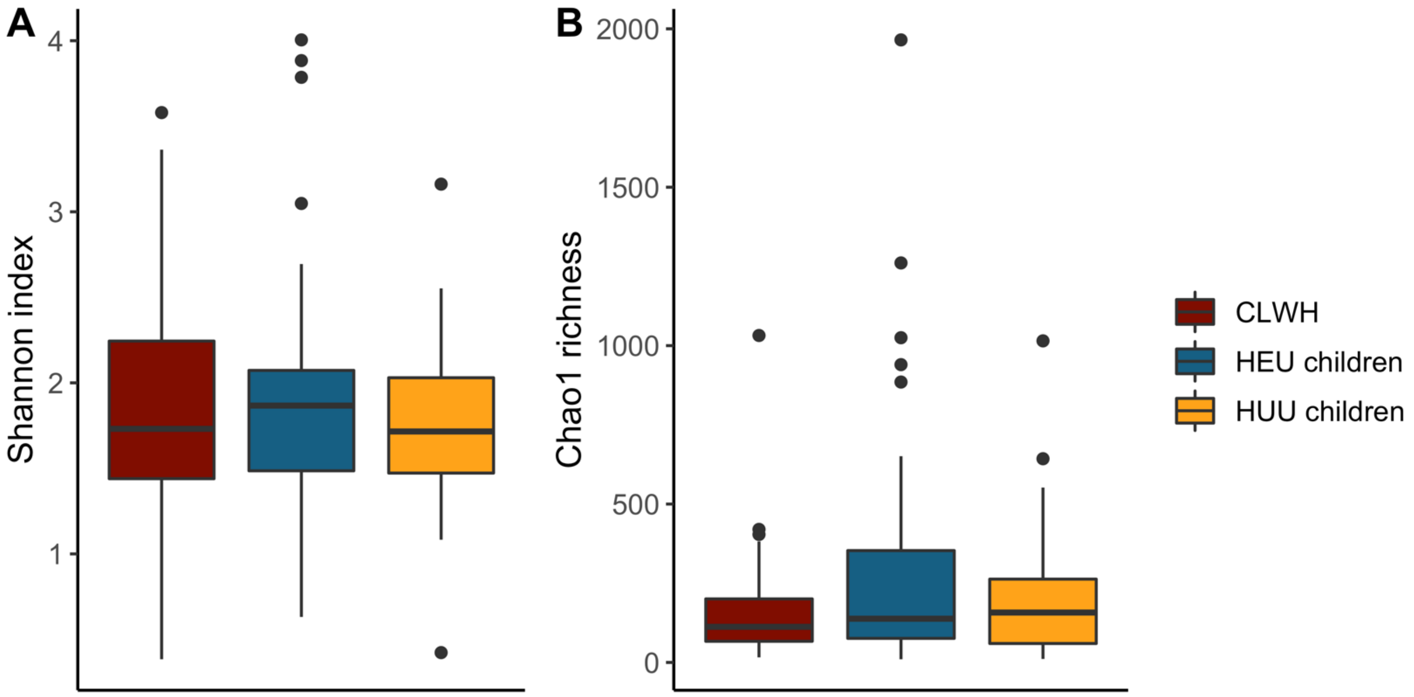

**Figure S3. Composition of the nasopharyngeal microbiome by HIV status among children with a history of breastfeeding in Botswana.** A. Principal components plot based on Euclidean distances showing distinct nasopharyngeal microbiome composition among CLWH (n=21) compared to HEU (n=16) and HUU children (n=47) with a history of breastfeeding in Botswana (PERMANOVA: $p=0{\cdot}007$, $R^2=0{\cdot}074$). B. Relative abundances of the ten most abundant bacterial species by HIV status in nasopharyngeal samples from children with a history of breastfeeding in Botswana.

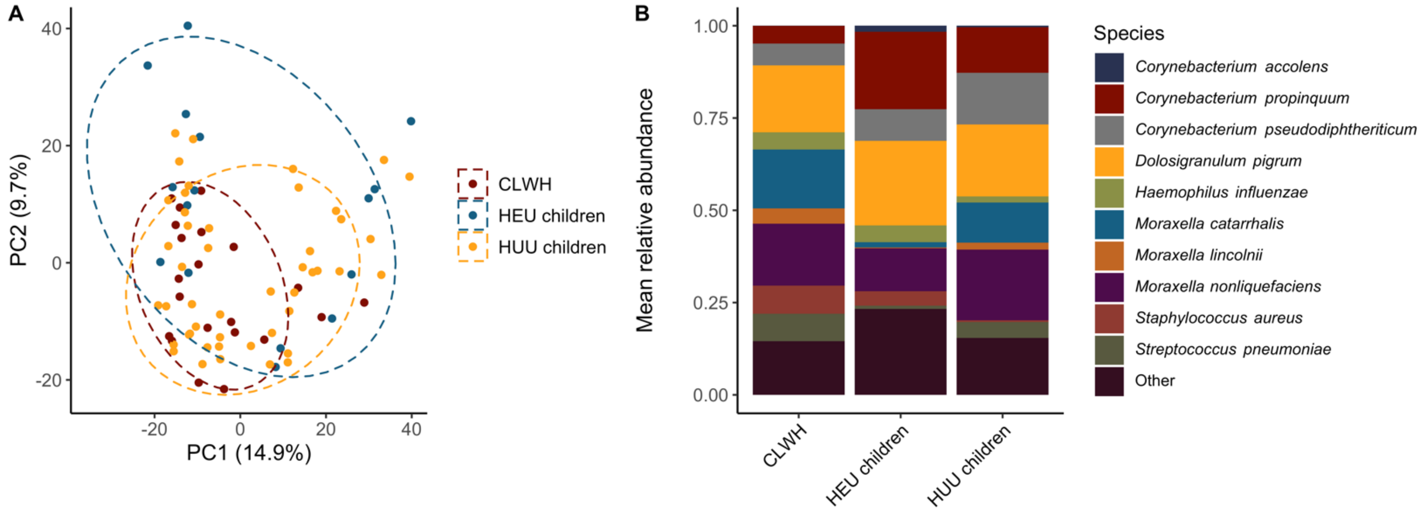

**Figure S4. Composition of the nasopharyngeal microbiome among sibling pairs.** Relative abundances of the ten most abundant bacterial species in nasopharyngeal samples from CLWH and their HEU siblings in Botswana (n=18). CLWH, children living with HIV. HEU, HIV-exposed, uninfected.

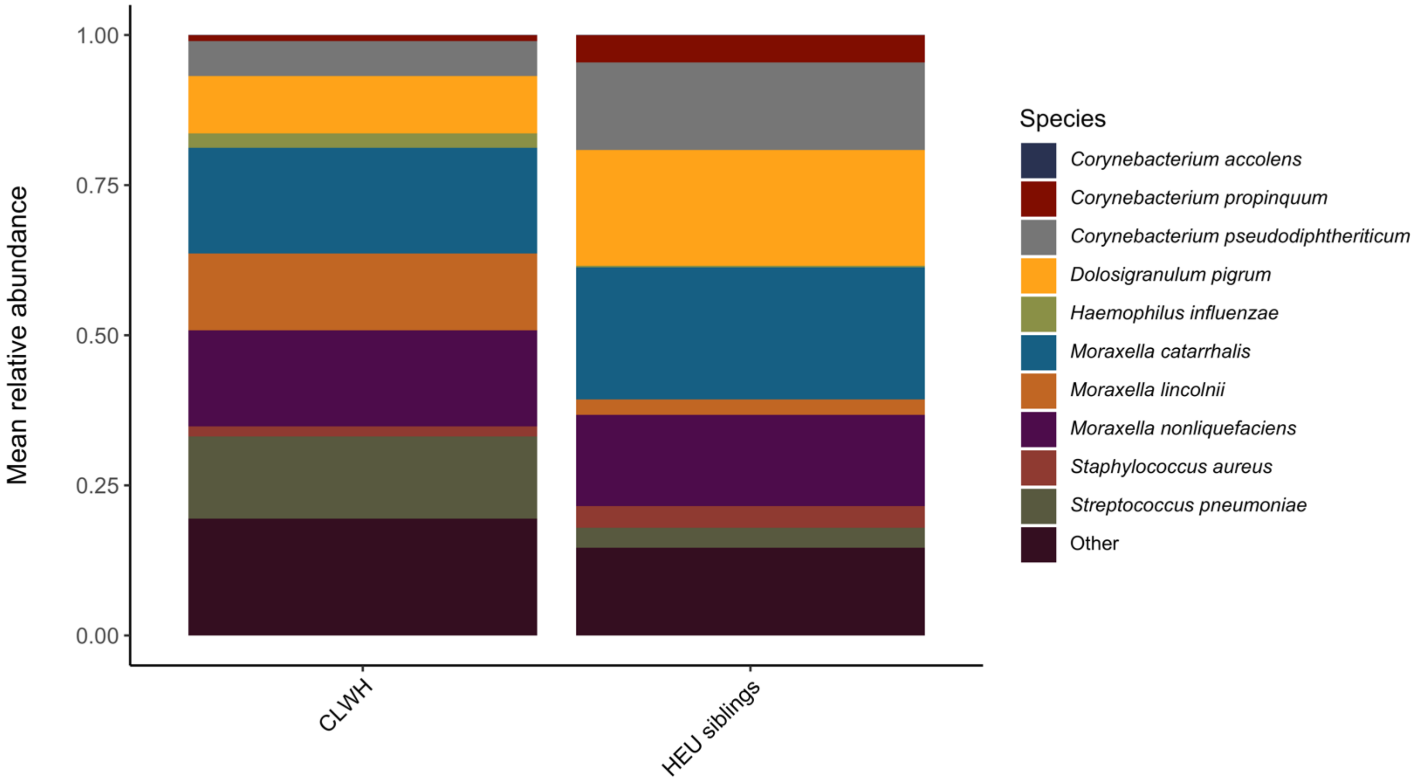

**Figure S5. Associations between HIV-associated factors and the composition of the microbiome among children in Botswana**. Linear discriminant effect size (LEfSe) was used to evaluate associations between HIV-related factors and the relative abundances of bacterial species within the nasopharyngeal microbiome. The coefficients from these models, which correspond to the relative effect sizes of associations, are shown for significant associations identified for (A) CD4+ % classification among CLWH (low vs normal, N=42), (B) TMP-SMX use among CLWH (n=42), and (C) immunosuppressed CLWH compared to HEU and HUU children (N=110). CLWH, children living with HIV; HEU, HIV-exposed, uninfected; HUU, HIV-unexposed, uninfected.

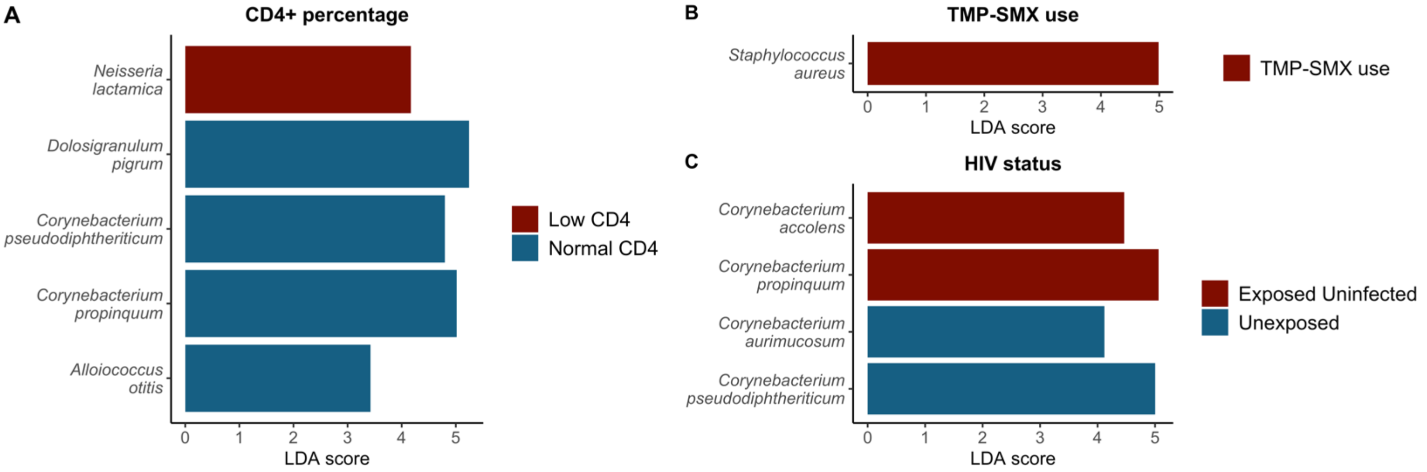

**Figure S6. Composition of the nasopharyngeal microbiome by HIV immune status among children in Botswana.** A. Principal components plot based on Euclidean distances showing distinct nasopharyngeal microbiome composition among immunosuppressed CLWH compared to HEU and HUU children in Botswana (n=110, PERMANOVA: $p = 0{\cdot}0030$, $R^2 = 0{\cdot}032$). B. Relative abundances of the ten most abundant bacterial species by HIV status in nasopharyngeal samples from children in Botswana, restricted to CLWH with a CD4+ cell percentage <25 (n=110). C. Principal components plot based on Euclidean distances showing no distinct nasopharyngeal microbiome composition among immunocompetent CLWH compared to HEU and HUU children in Botswana (n=130, PERMANOVA: $p=0{\cdot}15$, $R^2 = 0{\cdot}018$). D. Relative abundances of the ten most abundant bacterial species by HIV status in nasopharyngeal samples from children in Botswana, restricted to CLWH with a CD4+ cell percentage $\geq$25 (n=130).

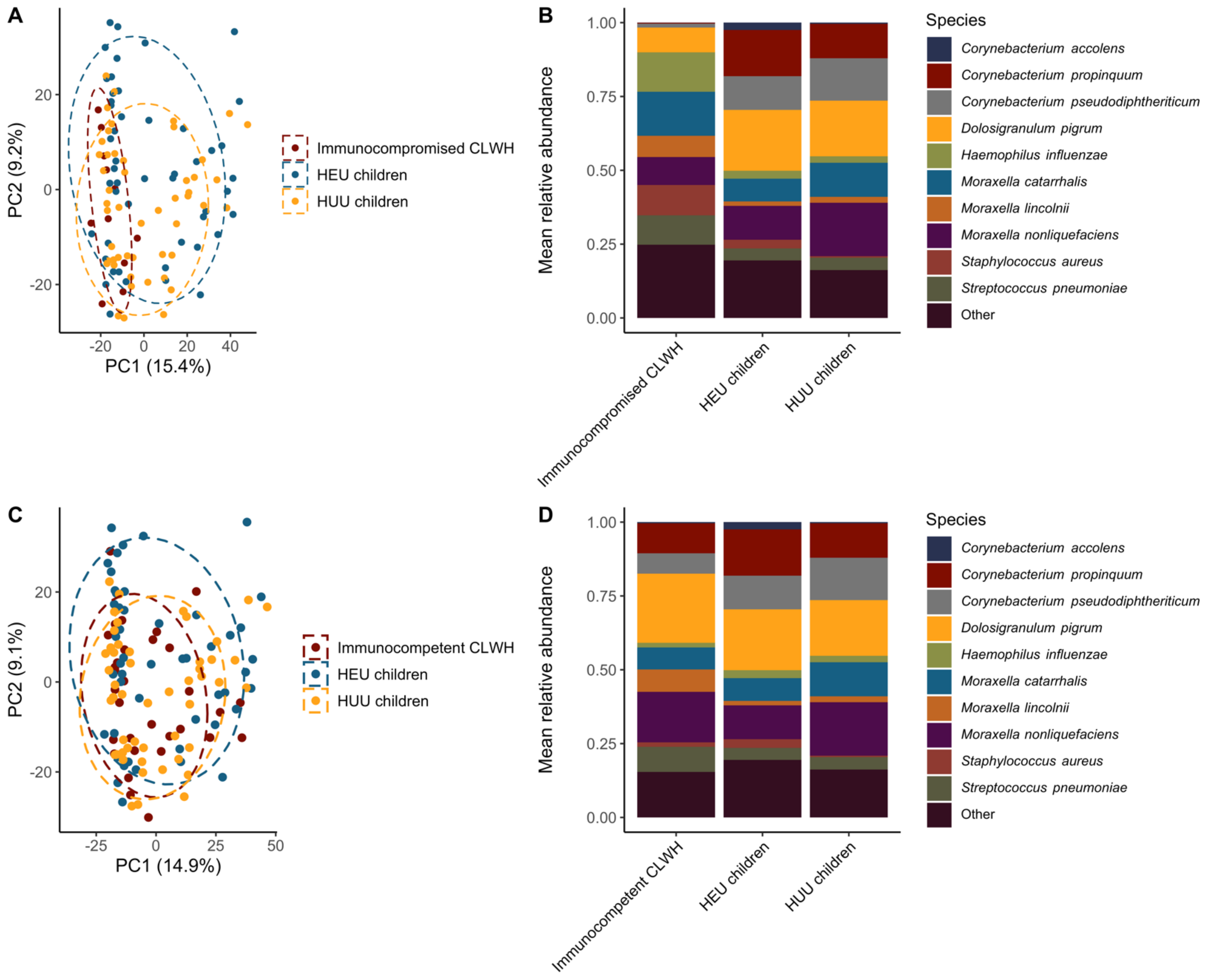

**Figure S7. Co-occurrence plot demonstrating associations between bacterial species in the nasopharyngeal microbiome.** Interspecies correlations were generated using SparCC. Each node represents a species. Pathogens of interest (*S. pneumoniae*, *S. aureus*) are colored green, while species differentially abundant by HIV status or HIV-associated immunosuppression are colored orange. Edges between nodes represent significant correlations. Blue edges represent positive correlations, red edges represent negative correlations, and the shade of red or blue represents the strength of the correlation coefficient. Only associations that passed false discovery rate correction are shown.

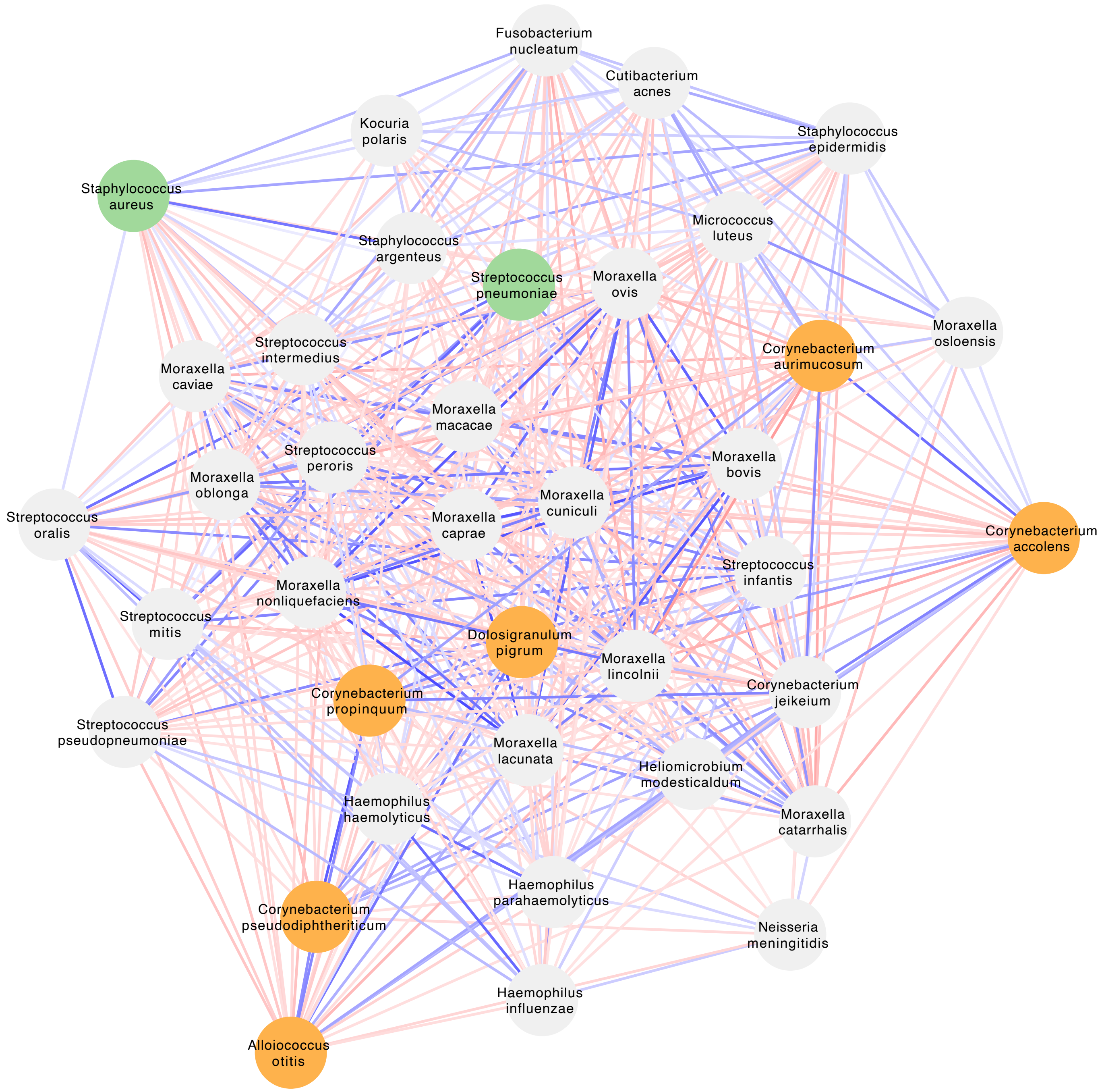
